# Diagnostic accuracy of the plasma ALZpath pTau217 immunoassay to identify Alzheimer’s disease pathology

**DOI:** 10.1101/2023.07.11.23292493

**Authors:** Nicholas J. Ashton, Wagner S. Brum, Guglielmo Di Molfetta, Andrea L. Benedet, Burak Arslan, Erin Jonatis, Rebecca E. Langhough, Karly Cody, Rachael Wilson, Cynthia M. Carlsson, Eugeen Vanmechelen, Laia Montoliu-Gaya, Juan Lantero-Rodriguez, Nesrine Rahmouni, Cecile Tissot, Jenna Stevenson, Stijn Servaes, Joseph Therriault, Tharick Pascoal, Alberto Lleó, Daniel Alcolea, Juan Fortea, Pedro Rosa-Neto, Sterling Johnson, Andreas Jeromin, Kaj Blennow, Henrik Zetterberg

**Author notes:** Corresponding author: Dr. Nicholas J. Ashton, Ph.D., Institute of neuroscience and physiology Dept. Psychiatry and Neurochemistry, Sahlgrenska Academy at Gothenburg University Mölndal Hospital, Hus V3, 43180 Mölndal, Sweden. Nicholas J. Ashton and Wagner S. Brum contribute equally as 1^st^ authors.

## Abstract

**Importance:** Phosphorylated tau (pTau) is a specific blood biomarker for Alzheimer’s disease (AD) pathology, with pTau217 considered to have the most utility. However, availability of pTau217 tests for research and clinical use has been limited. Expanding access to this highly accurate AD biomarker is crucial for wider evaluation and implementation of AD blood tests.

**Objective:** To determine the utility of a novel and commercially available Single molecule array (Simoa) for plasma pTau217 (ALZpath) to detect AD pathology. To evaluate references ranges for abnormal Aβ across three selected cohorts.

**Design, Setting, Participants:** Three single-centre observational cohorts were involved in the study: Translational Biomarkers in Aging and Dementia (TRIAD), Wisconsin Registry for Alzheimer’s Prevention (WRAP), and Sant Pau Initiative on Neurodegeneration (SPIN). MRI, Aβ-PET, and tau-PET data were available for TRIAD and WRAP, while CSF biomarkers were additionally measured in a subset of TRIAD and SPIN. Plasma measurements of pTau181, pTau217 (ALZpath), pTau231, Aβ42/40, GFAP, and NfL, were available for all cohorts. Longitudinal blood biomarker data spanning 3 years for TRIAD and 8 years for WRAP were included.

**Exposures:** MRI, Aβ-PET, tau-PET, CSF biomarkers (Aβ42/40 and pTau immunoassays) and plasma pTau217 (ALZpath Simoa).

**Main Outcomes and Measures:** The accuracy of plasma pTau217 for detecting abnormal amyloid and tau pathology. Longitudinal pTau217 change according to baseline pathology status.

**Results:** The study included 786 participants (mean [SD] age, 66.3 [9.7] years; 504 females [64.1%]) were included in the study. High accuracy was observed in identifying elevated Aβ (AUC, 0.92-0.96; 95%CI 0.89-0.99) and tau pathology (AUC, 0.93-0.97; 95%CI 0.84-0.99) across all cohorts. These accuracies were significantly higher than other plasma biomarker combinations and comparable to CSF biomarkers. The detection of abnormal Aβ pathology using binary or three-range references yielded reproducible results. Longitudinally, plasma pTau217 showed an annual increase only in Aβ-positive individuals, with the highest increase observed in those with tau-positivity.

**Conclusions and Relevance:** The ALZpath plasma pTau217 Simoa assay accurately identifies biological AD, comparable to CSF biomarkers, with reproducible cut-offs across cohorts. It detects longitudinal changes, including at the preclinical stage, and is the first widely available, accessible, and scalable blood test for pTau217 detection.

**Key Points:** *Question:* What are the capabilities of the ALZpath plasma pTau217 Single molecule array (Simoa) to identify Alzheimer’s disease (AD) pathophysiology?

*Findings:* ALZpath pTau217 showed similar accuracies to cerebrospinal fluid biomarkers in identifying abnormal Aβ and tau pathologies. Calculated reference ranges for detecting abnormal Aβ were consistent across three cohorts. Over 8 years, the largest change of pTau217 was in individuals positive for both Aβ and tau.

*Meaning:* These results demonstrate the high-performance of the ALZpath plasma pTau217 Simoa in identifying AD-type pathophysiology. The wider availability of high-performing assays will expedite the use of blood biomarkers in clinical settings and benefit the research community.

## Introduction

In Alzheimer’s disease (AD), blood biomarkers have emerged as scalable tools for clinical evaluation, trial recruitment, and disease monitoring^1^. The anticipated implementation of such tests would substantially reduce the reliance on confirmatory evaluations with cerebrospinal fluid (CSF) or positron emission tomography (PET) scans in specialized centers^2^. An accurate and robust blood-based biomarker for AD pathology would enable a more comprehensive biological assessment of cognitive impairment in settings where advanced testing is limited *e.g.,* primary care. Therefore, the aim of a blood biomarker is to enhance an early and precise AD diagnosis, leading to improved patient management and timely access to disease-modifying therapies.

Phosphorylated tau (pTau) is the leading candidate among novel AD blood tests, demonstrating superior diagnostic accuracy and disease specificity compared to other proposed blood biomarkers^3, 4^. The Aβ42/40 ratio, validated for use in CSF^5^, has limitations in blood due to its small fold-change between amyloid PET-positive and amyloid PET-negative individuals^6, 7^. Consequently, plasma Aβ42/40 lacks the clinical robustness required for routine clinical testing^8, 9^. In contrast, high-performing pTau blood tests exhibit a 100-400% increase in AD patients^10^, with increases occurring concurrently with extracellular Aβ plaque pathology, an AD hallmark feature. This relationship is observed across the AD continuum, including pre-symptomatic changes in sporadic and familial AD^11^, as well as in individuals with Down syndrome who have a genetically determined form of AD^12–14^. Furthermore, certain pTau species are associated with neurofibrillary tangle pathology, the secondary AD pathological hallmark, as indexed by tau-PET imaging and neuropathological examination^15–17^. Thus, pTau is regarded as the primary blood biomarker for AD pathology throughout all stages of the disease.

Among proposed blood pTau biomarkers^18–21^, phosphorylated tau at Threonine 217 (pTau217) has consistently shown high-performance in differentiating AD from other neurodegenerative disorders^10, 22^ and in detecting AD pathology in MCI patients^22^. Notably, pTau217 exhibits larger fold-changes compared to pTau181 and pTau231^10^, often achieving high discrimination, with areas under the curve (AUC) exceeding 90%^19, 23^. Additionally, pTau217 demonstrates a unique longitudinal trajectory in amyloid-positive individuals, showing increases over time significantly associated with worsening cortical atrophy and declining cognitive performance^24, 25^.

With the impending implementation of anti-Aβ therapies in dementia management, validated blood biomarkers are urgently needed to guide timely treatment decisions. These biomarkers should be accessible and compatible with analytical platforms used in diagnostic laboratories of all sizes and locations. While pTau217 has shown promise as a diagnostic tool for AD, its widespread evaluation has been hindered by limited availability of commercially available assays. This study aims to address this gap by assessing the diagnostic utility of ALZpath pTau217, a commercially available single molecule array (Simoa), and evaluating reference ranges for AD pathology.

## Methods

### Participants & ethics

This study included participants from three observational cohorts: the Translational Biomarkers in Aging and Dementia (TRIAD), Wisconsin Registry for Alzheimer’s Prevention (WRAP), and the Sant Pau Initiative on Neurodegeneration (SPIN). Informed consent was obtained from all participants, and the studies were approved by the relevant ethics boards (**Supplementary Methods**). TRIAD included 268 participants categorized as cognitively unimpaired (CU; n [%]: 134[50%]), mild cognitive impairment (MCI; 63[23.5%]), AD dementia (46[17.2%]) and non-AD dementia (24[9.0%]). The WRAP study^26^ included data on 323 participants, predominantly CU at the first plasma sample collection (CU: 309[95.6%]; MCI: 12[3.7%]; AD dementia: 2[0.6%]). The SPIN cohort^27^ included 195 participants consisting of CU controls (82[42.1%]), MCI due to AD (72[36.7%]), and AD dementia (41[21.0%]). Diagnosis was based on internationally recognized clinical criteria, and control participants had normal cognitive scores on standard neuropsychological evaluations.

A subset of patients with longitudinal follow-up consisted of 392 participants from TRIAD and WRAP (**Supplementary Table 1**). These included participants classified into three groups: amyloid- and tau-negative (A-T-; n=297), amyloid-positive and tau-negative (A+T-; n=66), and amyloid-positive and tau-positive (A+T+; n=29). In WRAP, the median number of samples collected per patient was three, over a mean (SD) of 5.22 (1.41) years. In TRIAD, median samples per patient was two, collected over a mean of 1.90 (0.61) years.

### Imaging

Detailed imaging methods for TRIAD, WRAP and SPIN are found in the **Supplementary Methods**. In TRIAD, Aβ-and tau-PET were determined by [^18^F]-AZD4694^28^ and [^18^F]-MK6240^29^, respectively. In WRAP, PET measures were determined by [^11^C]-PiB^30^ and [^18^F]-MK6240^31, 32^. In SPIN, Aβ-PET was determined by [^18^F]-florbetapir or [^18^F]-flutemetamol in a subset of participants. Aβ-PET-positivity was standardised across cohorts as a Centiloid value of >24 (SUVR>1.55 for [^18^F]-AZD4694 or DVR>1.2 for [^11^C]-PiB). Tau-positivity with [^18^F]-MK6240 was defined as meta-temporal SUVR>1.24 for TRIAD^33^ and as entorhinal SUVR>1.3 in WRAP. In WRAP, the mean time differences between first plasma sample and scan were 0.94 years for Aβ-PET and 2.27 years for tau-PET; in TRIAD, this was 0.44 years for Aβ-PET and 0.42 years for tau-PET.

### Cerebrospinal fluid biomarkers

CSF sample collection procedures were similar across cohorts. Samples were collected with needle drip technique into polypropylene tubes followed by 2200g centrifugation for 10 minutes. Samples were then distributed into 0.25–1 mL aliquots in polypropylene vials (Fisher Scientific Inc. Catalog #3741-WP1D-BR or Sarstedt 2mL #72.694.007) and stored at -80 °C until biochemical analyses. In TRIAD and SPIN, LUMIPULSE G1200 or G600II were used to quantify CSF Aβ42, Aβ40 and pTau181^34, 35^. Additionally, CSF pTau217 was quantified by an *in-house* single molecule array (Simoa) developed at the University of Gothenburg^36^. A novel Simoa for CSF pTau205 was measured in TRIAD only. For WRAP, CSF Aβ42, Aβ40, pTau181 were measured using the Roche NeuroToolKit^37^.

### Plasma biomarkers

Plasma samples from TRIAD, WRAP and SPIN were analyzed at the Department of Psychiatry and Neurochemistry, University of Gothenburg. Plasma Aβ42/40, GFAP and NfL were quantified using the commercial Neurology 4-plex E kit (#103670, Quanterix). Plasma pTau231 and pTau181 were analysed using *in-house* Simoa assays developed at the University of Gothenburg^18^ ^38^, except in WRAP where plasma pTau181 was quantified by the commercial Advantage V2.1 kit (#104111, Quanterix) ^24^.

### ALZpath pTau217 assay

ALZpath has developed a novel commercial plasma-based Simoa assay for pTau217, utilizing a proprietary monoclonal pTau217 specific capture antibody, an N-terminal detector antibody and a peptide calibrator. It has been validated as a *fit-for-purpose* assay, with a limit of detection of 0.0052-0.0074 pg/mL, a functional lower limit of quantification of 0.06 pg/mL (based on precision profile with CV_%_<20), a dynamic range of 0.007–30 pg/mL (minimal required dilution of 3). The spike recovery of for the endogenous analyte was 80%, and intra-and inter-run precision was 0.5–13% and 9.2–15.7%, respectively. Here, the assay demonstrated good repeatability (TRIAD, 4–5.4%; WRAP, 6.6–8.7%; SPIN, 4.1–5.3%) and intermediate precision (TRIAD, 7.1–8.1%; WRAP, 3.5–10.7%; SPIN, 7.1–8-1%) using three internal plasma quality controls samples from the University of Gothenburg and two quality controls provided via the ALZpath assay. Summary of precision results can be found in **Supplementary Table 2**.

### Statistical analysis

Between-group comparisons of plasma pTau217 levels were conducted using linear models, adjusting for age and sex. Discriminative ability of plasma pTau217 for Aβ- and tau-PET-positivity, and other outcomes, was assessed using receiver operating characteristics area under the curve (AUC) and was compared to that of other established biomarkers with the DeLong test, when applicable. Correlations were always evaluated using Spearman’s rho. Two reference range strategies were evaluated for interpreting plasma pTau217 results. As conventionally done, a binary reference-point for Aβ-PET-positivity was derived based on the Youden index. Alternatively, a strategy comprising a lower reference-point to rule out AD (95% sensitivity) and a higher reference-point to rule in AD (95% specificity). In both strategies, we evaluated the concordance of a negative pTau217 result with Aβ-PET negativity (negative percent agreement; NPA), and the concordance of a positive plasma p-tau217 with Aβ-PET-positivity (positive percent agreement; PPA), as well as the overall percent agreement (OPA). In the latter strategy, individuals with pTau217 levels between the reference-point were classified as intermediate-risk and would constitute the population referred to confirmatory testing^38^. In the two cohorts with serial pTau217 data, we evaluated the longitudinal trajectories of plasma pTau217 in CU and MCI participants according to their PET-defined amyloid and tau status. We used linear mixed effects models with plasma pTau217 as the response variable, including as predictors time (since first plasma collection), AT status, age at first plasma collection, years of education, sex, and cognitive status at first visit, as well as an interaction between AT status and time. The model contained random intercepts and random slopes for each participant, and time was modelled as a continuous variable. Post-hoc pairwise contrasts were conducted to compare the slopes for group-by-time interactions. All analyses were performed using R version 4.2.2 (https://www.r-project.org/), with a two-sided alpha of 0.05. Reported results include 95% confidence intervals when applicable.

## Results

### Participant characteristics

A total of 786 participants (mean [SD] age, 66.3 [9.7] years; *n* [%] 504 females [64.1%]) were included in the study. The TRIAD subsample included 268 participants (69.4 [7.8] years; 167 [62.3%] females). The WRAP cohort included 323 participants (65.3 [6.9] years; 217 [67.2%] females), predominantly CU. The SPIN cohort included 195 participants (63.5 [13.8] years; 120 [61.5%] females). All included participants had confirmatory amyloid status (TRIAD and WRAP: Aβ-PET; SPIN: CSF Aβ42/Aβ40), and the vast majority (716 [91.1%]) also had information on tau status (TRIAD and WRAP: tau-PET; SPIN: CSF p-tau181), as described in **Table 1** alongside demographic and clinical information for all cross-sectional analyses. **Supplementary Table 1** describes the TRIAD (n=132) and WRAP (n=260) longitudinal subsets.

**Table 1.** Cross-sectional demographics of WRAP, TRIAD, and SPIN cohorts. Data are mean (SD) or n (%). In WRAP and TRIAD, AT status was defined with amyloid and tau-PET. In TRIAD, all participants had available Aβ-PET data (100%), while tau-PET was not available for 57 (17.6%) participants. In WRAP all participants had available Aβ-PET data (100%), and tau-PET was not available for 2 (0.7%) participants. In SPIN, all participants had data for amyloid status, which was determined with CSF Aβ42/Aβ40 in 159 (71.5%) participants or with Aβ-PET in 36 (18.5%) participants. In SPIN, tau status was defined with CSF p-tau181 and was not available for 11 participants (5.6%). Abbreviations: SD, standard deviation; TRIAD, Translational Biomarkers in Aging and Dementia; WRAP, Wisconsin Registry for Alzheimer’s Prevention; SPIN, Sant Pau Initiative on Neurodegeneration; MMSE, mini-metal state examination; CU, cognitively unimpaired; CI, cognitively impaired; A-T-, amyloid-negative and tau-negative; AT-, amyloid-positive and tau-negative; A-T+, amyloid-positive and tau-positive.

|  | WRAP<br>(n=323) | TRIAD<br>(n=268) | SPIN<br>(n=195) |
| --- | --- | --- | --- |
| Age, years, mean (SD) | 65.3 (6.91) | 69.4 (7.90) | 63.5 (13.8) |
| Female, n (%) | 217 (67.2%) | 167 (62.3%) | 120 (61.5%) |
| *APOE* ε4 carriers, n (%) | 121 (37.5%) | 96 (35.8%) | 81 (41.5%) |
| MMSE score, mean (SD) | 29.2 (1.23) | 27.0 (4.72) | 26.4 (4.19) |
| Baseline clinical diagnosis, n (%) |  |  |  |
| CU | 309 (95.7%) | 134 (50.0%) | 82 (42.1%) |
| CI | 14 (4.3%) | 134 (50.0%) | 113 (57.9%) |
| Years of education,<br>mean (SD) | 16.1 (2.62) | 15.0 (3.57) | 13.4 (5.12) |
| AT status, n (%) |  |  |  |
| A-T- | 209 (78.9%) | 146 (55.3%) | 75 (41.2%) |
| A+T- | 38 (14.3%) | 65 (24.6%) | 6 (3.3%) |
| A+T+ | 18 (6.8%) | 53 (20.1%) | 101 (55.5%) |
| A-T+ | 1 (0.3%) | 2 (0.7%) | 2 (1.0%) |
| Missing | 57 (17.6%) | 2 (0.7%) | 11 (5.6%) |
| Plasma pTau217, pg/mL, mean (SD) | 0.466 (0.362) | 0.636 (0.648) | 0.977 (0.766) |

### Levels of ALZpath pTau217 by amyloid “A” and tau “T” status

When stratified by amyloid (“A”) and tau (“T”) status, regardless of clinical diagnosis, plasma pTau217 significantly increased in a stepwise manner in all cohorts (**Figure 1**), with highest levels in the A+T+ group. Mean pTau217 concentrations (pg/mL) for A-T-(mean [SD] TRIAD, 0.26 [0.13]; WRAP, 0.35 [0.15]; SPIN, 0.32 [0.11]), A+T-(TRIAD, 0.75 [0.63]; WRAP, 0.72 [0.30]; SPIN, 0.91 [0.47]), and A+T+ (TRIAD, 1.48 [0.65]; WRAP, 1.41 [0.70]; SPIN, 1.50 [0.70]), were remarkably similar across all three cohorts. This was also observed when stratifying by amyloid status alone (A-, TRIAD, 0.28 [0.21], WRAP, 0.35 [0.14]; SPIN, 0.38 [0.29]); A+, TRIAD, 1.08 [0.72], WRAP, 0.94 [0.54], SPIN, 1.43 [0.70] (**Supplementary Figure 1**).

**Figure 1.**
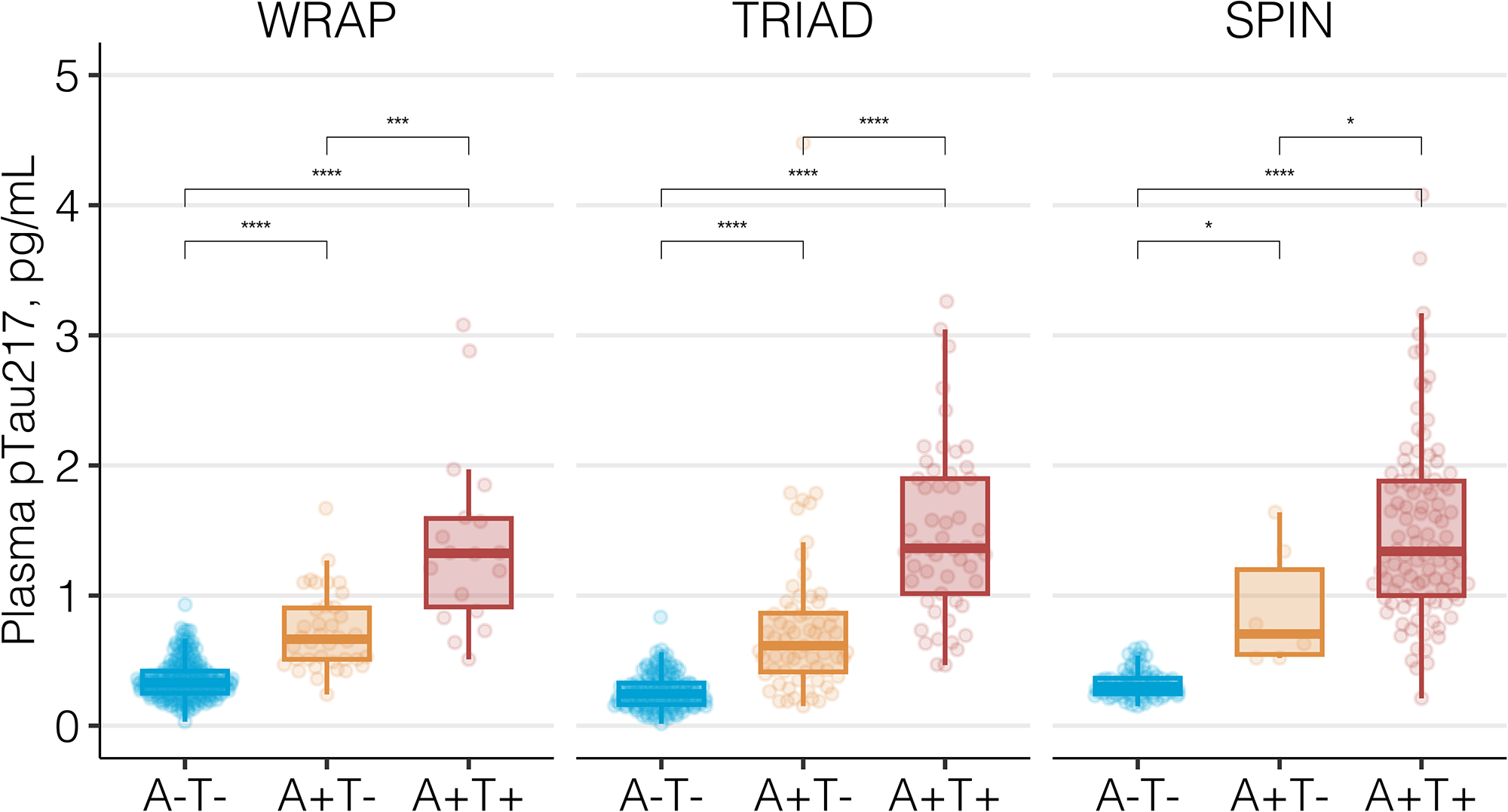
Plasma ALZpath pTau217 levels according to amyloid and tau profiles. Boxplots show the distribution of ALZpath pTau217 concentrations by AT profile for the WRAP, TRIAD, and SPIN cohorts. For WRAP and TRIAD, Aβ (“A”) and tau (“T”) were indexed by PET. In SPIN, “A” was indexed by CSF Aβ42/40 and “T” by pTau181. All comparison p-values, obtained from pairwise contrasts from linear models adjusted for age and sex were <0.0001 (****), whereas in the SPIN cohort two comparisons showed p<05 (p<0.05 = *; A-T-vs A+T-: p=0.018; A+T-vs A+T+: p=0.032).

### ALZpath pTau217 discriminates abnormal Aβ and tau pathologies with high accuracy

ALZpath plasma pTau217 demonstrated high accuracy in predicting abnormal Aβ-PET signal (**Figure 2A**) in TRIAD (AUC, 0.92; 95%CI, 0.92-0.96) and WRAP (AUC, 0.93; 95%CI, 0.90-0.97). In SPIN, ALZpath pTau217 also had high accuracy in predicting abnormal CSF Aβ42/40 (AUC, 0.96; 95%CI, 0.92-0.99; **Figure 2A**). There was equally high accuracy when Aβ-PET status was determined by visual read (**Supplementary Figure 2)**, and cases classified as “intermediate” or “discordant” by visual read had also intermediate median pTau217 levels (**Supplementary Figure 2**). ALZpath pTau217 also exhibited high accuracy for predicting abnormal tau (**Figure 2B**) in TRIAD (AUC, 0.95; 95%CI, 0.92-0.97) and WRAP (AUC, 0.93; 95%CI, 0.84-0.98). In SPIN,

**Figure 2.**
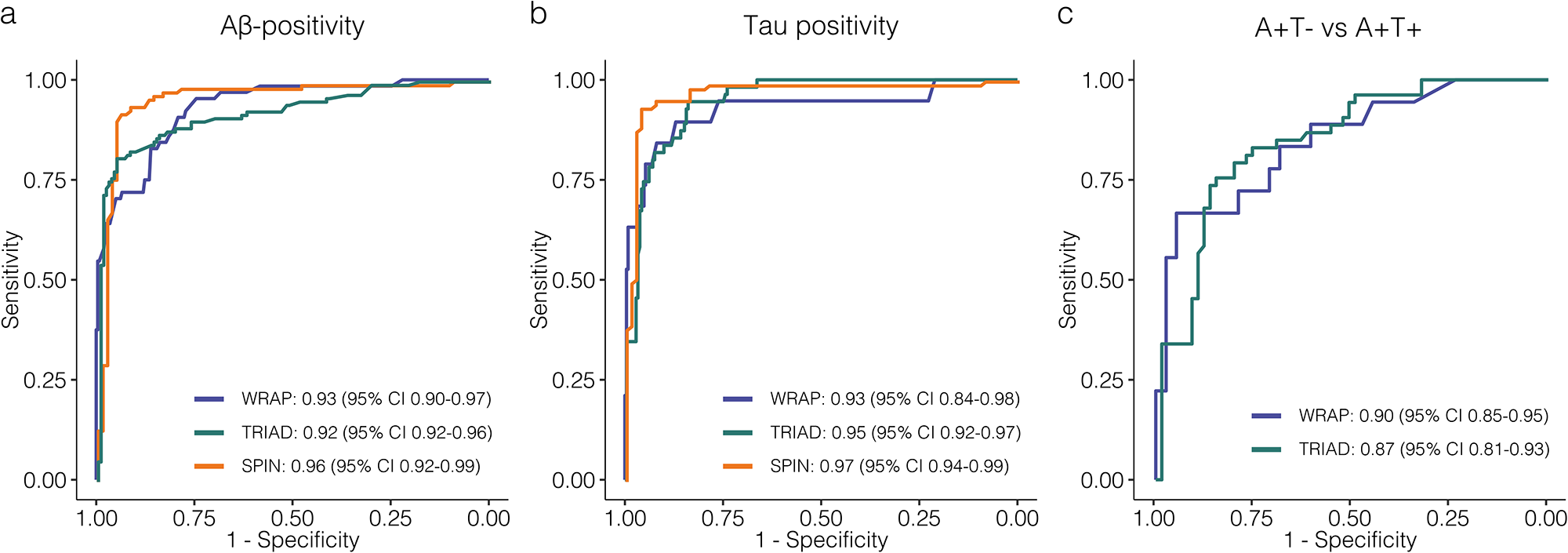
Accuracy of ALZpath pTau217 for decting A**β**-positivity, tau-positivity and discriminating A+T-from A+T+ individuals. Receiver operating characteristics (ROC) curves for ALZpath pTau217 to detecting Aβ-positivity (A), tau-positivity (B) and to differentiate Aβ-positive and tau-positive individuals (A+T+) from Aβ-positive and tau-negative (A+T-) (C). Solid lines vary in color according to each cohort, as depicted in the legend. For each ROC curve, the area under the curve (AUC) is reported alongside 95% confidence intervals (CI).

ALZpath pTau217 had high accuracy for abnormal CSF pTau181 (AUC, 0.97; 95%CI, 0.94-0.99). Promisingly, ALZpath pTau217 could identify abnormal tau among amyloid-positive (A+T-vs A+T+) participants in TRIAD (AUC, 0.87; 95%CI, 0.81-0.93) and WRAP (AUC, 0.90; 95%CI, 0.85-0.95) (**Figure 2C**). Moreover, we observed a gradual increase of plasma pTau217 across tau-PET-defined Braak stages in TRIAD (**Supplementary Figure 3**). The highest levels of pTau217 were observed in Braak V-VI (mean [SD], 1.55 [0.70]) which was significantly higher than Braak III-IV (0.92 [0.46]; *P*<0.001), Braak I-II (0.49 [0.35]; *P*<0.001) and Braak 0 (0.30 [0.41]; *P*<0.001).

### ALZpath pTau217 is comparable to imaging and CSF biomarkers in identifying AD pathology

Next, we compared the performance of ALZpath plasma pTau217 against other CSF and imaging modalities for predicting abnormal Aβ-PET (**Figure 3A–C**) and tau-PET (**Figure 3D–E**). This analysis included the maximum number participants within each biomarker modality.

**Figure 3.**
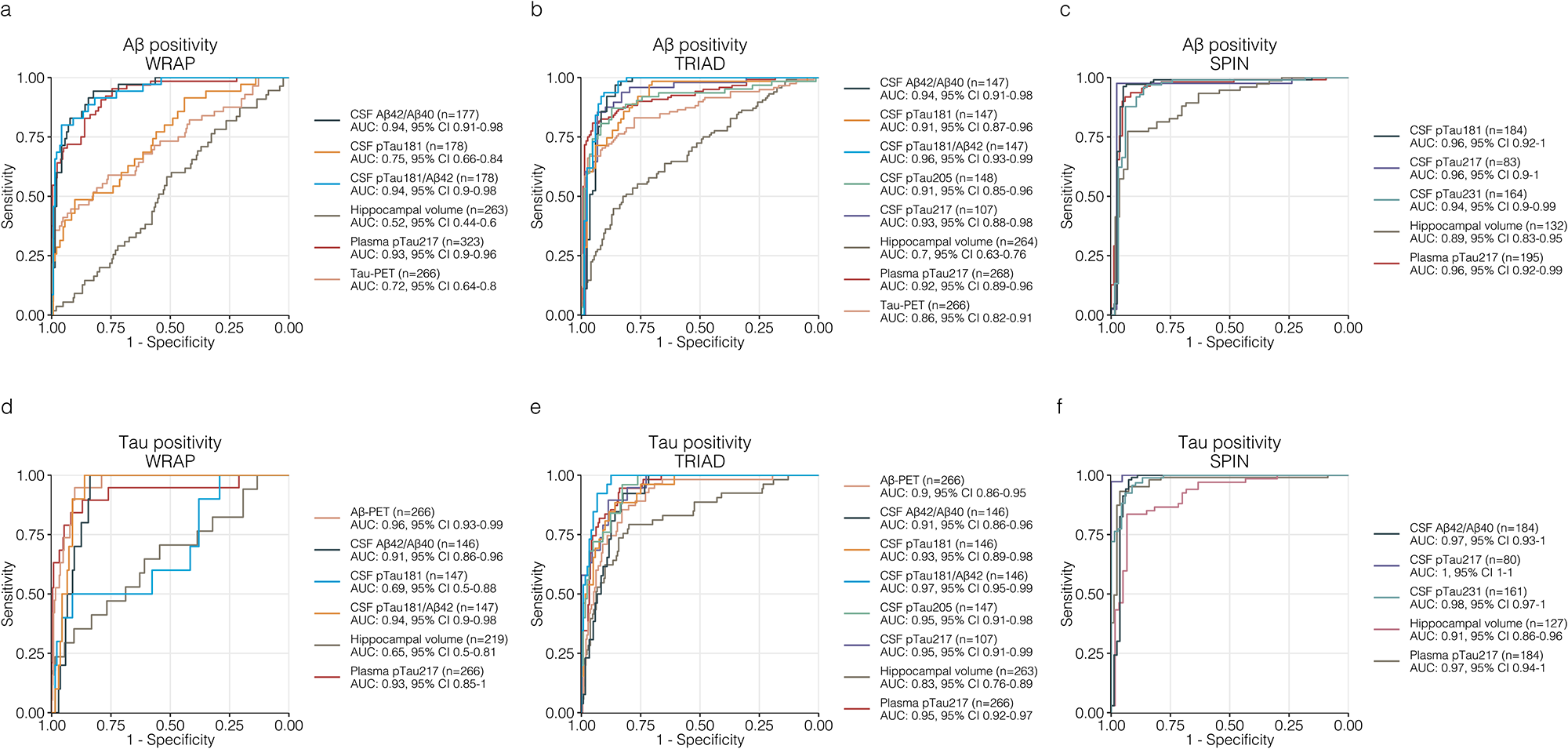
Plasma ALZpath pTau217 demonstrates similar or superior diagnostic accuracy for Aβ and tau pathologies compared to established CSF and PET biomarkers. Receiver operating characteristics curves for detecting Aβ (A-C) and tau-positivity (D-F) for ALZpath pTau217, CSF and imaging biomarkers. Each panel corresponds to accuracies for the same outcome at each cohort (WRAP: A, D; TRIAD: B, E; SPIN: C, F). Solid lines represent the ROC curves for each biomarker, with colors corresponding to a specific biomarker across cohorts, as indicated in the figure legend at each panel. The legend indicates the area under the curve (AUC) for each biomarker, alongside 95% confidence intervals (CI). The maximum number of observations with complete data for each biomarker and outcome were used, and the specific number for each biomarker is also detailed in the figure legend. This approach, in which no data is excluded, yielded similar results to AUC comparisons within a reduced subset including the same subjects with complete data for all biomarkers, reported in the Supplement.

In WRAP, in determining abnormal Aβ-PET, plasma pTau217 outperformed hippocampal atrophy (AUC, 0.52; 95%CI, 0.44-0.60; *P*<0.001), tau-PET (AUC, 0.72; 95%CI, 0.64-0.80; *P*<0.001) and CSF pTau181 (AUC, 0.75; 95%CI, 0.66-0.84; *P*<0.01), but did not differ significantly from CSF Aβ42/40 or CSF pTau181/Aβ42 (all p>0.05, **Figure 3A**). Similar findings were observed in TRIAD, where plasma pTau217 outperformed hippocampal atrophy (AUC, 0.70; 95%CI, 0.63-0.76; *P*<0.0001) and tau-PET (AUC, 0.86; 95%CI, 0.82-0.91; *P*=0.05) for detecting abnormal Aβ pathology but did not significantly differ from various CSF biomarkers (all p>0.05, **Figure 3B**). In SPIN, plasma pTau217 outperformed hippocampal volume (AUC, 0.89; 95%CI, 0.83-0.95; *P*=0.04) and was comparable to CSF biomarkers (all p>0.05, **Figure 3C**).

In predicting abnormal tau-PET burden (**Figure 3D–E**), plasma ALZpath pTau217 significantly outperformed hippocampal volume (AUC^WRAP^, 0.65; 95%CI, 0.50-0.81, *P*=0.01; AUC^TRIAD^, 0.83; 95%CI, 0.76-0.89; *P*=0.01; AUC^SPIN^, 0.91; 95%CI, 0.86-0.96, *P*=0.049). Plasma pTau217 significantly outperform CSF pTau181 in WRAP (AUC, 0.69; 95%CI, 0.66-0.84; *P*=0.02) but not in TRIAD. Plasma pTau217 outperformed Aβ-PET in TRIAD (AUC, 0.90; 95%CI, 0.86-0.95; *P*=0.04), while in WRAP they were comparable (AUC, 0.96; 95%CI, 0.93-0.99; *P*=0.35). Plasma pTau217 showed comparable performance to other measures, except for CSF pTau217 in SPIN, which was superior (*P*=0.04).

Additionally, we conducted comparisons in subsets only including participants with all modalities (WRAP: n=131; TRIAD: n=106; SPIN: n=41), finding no marked differences (**Supplementary Figure 4**). Plasma pTau217 also discriminated A+T+ from A+T-individuals comparably to CSF and imaging biomarkers (**Supplementary Figure 5**).

### ALZpath pTau217 outperformed other plasma biomarkers, alone or in combination, to identify AD pathology

Plasma pTau217 alone or pTau217 plus demographic variables (age, sex and APOE status) outperformed all other plasma biomarkers (pTau181, pTau231, Aβ42/40, GFAP and NfL), and their optimal combinations, for predicting both amyloid and tau status in all cohorts (**Supplementary Table 4; Supplementary Table 5; Supplementary Figure 6**). A minimal improvement in model metrics of goodness-of-fit (Akaike information criterion) was observed in pTau217 plus demographics but not in discriminatory performance.

### Correlations of ALZpath plasma pTau217 with Aβ-PET, tau-PET, and CSF biomarkers

Plasma pTau217 correlated strongly with Aβ-PET in WRAP (*Rho*=0.75, *P*<0.0001; **Supplementary Figure 7A**) and TRIAD (*Rho*=0.72, *P*<0.0001**; Supplementary Figure 7B**). In the subset of SPIN participants with Aβ-PET a similar association was found (*Rho* = 0.56, *P*<0.0001; **Supplementary Figure 7C**). Further, strong associations were found for tau-PET in WRAP (*Rho*=0.73, *P*<0.0001; **Supplementary Figure 7D**) and TRIAD (*Rho*=0.78, *P*<0.0001**; Supplementary Figure 7E**). Plasma pTau217 was also shown to be highly correlated with CSF pTau217 in TRIAD (*Rho* = 0.78, *P*<0.0001 **Supplementary Figure 8A)** and SPIN (*Rho*=0.88, *P*<0.0001 **Supplementary Figure 7B)**.

### Reference ranges for abnormal Aβ and tau pathologies based on ALZpath plasma pTau217

We first derived a binary reference-point for Aβ-positivity using the Youden index, derived in WRAP (>0.42 pg/mL, **Figure 4A, Supplementary Table 6**). This reference-point was cross-validated in TRIAD (Aβ-positivity based on PET) and SPIN (Aβ-positivity based on CSF Aβ42/40). Acknowledging the existence of an overlap in pTau217 levels, as observed with all high-preforming blood assays, lower (95% sensitivity, <0.4 pg/mL) and upper (95% specificity, >0.63 pg/mL) reference-points were derived in WRAP (**Figure 4B, Supplementary Table 7**). This approach improved the PPA (TRIAD: 97.7%; SPIN: 95.3%) while maintaining a similar NPA. The pair of reference-points created an “intermediate” zone (pTau217 levels between 0.4-0.63 pg/mL), which could in practice be referred to confirmatory testing with CSF or PET. The intermediate zone was largest in WRAP (22.9%), as expected due to lower Aβ-positivity prevalence, and smaller in TRIAD (15.8%) and SPIN (13.0%). A binary reference-point for tau-positivity is demonstrated in **Supplementary Table 8**.

**Figure 4.**
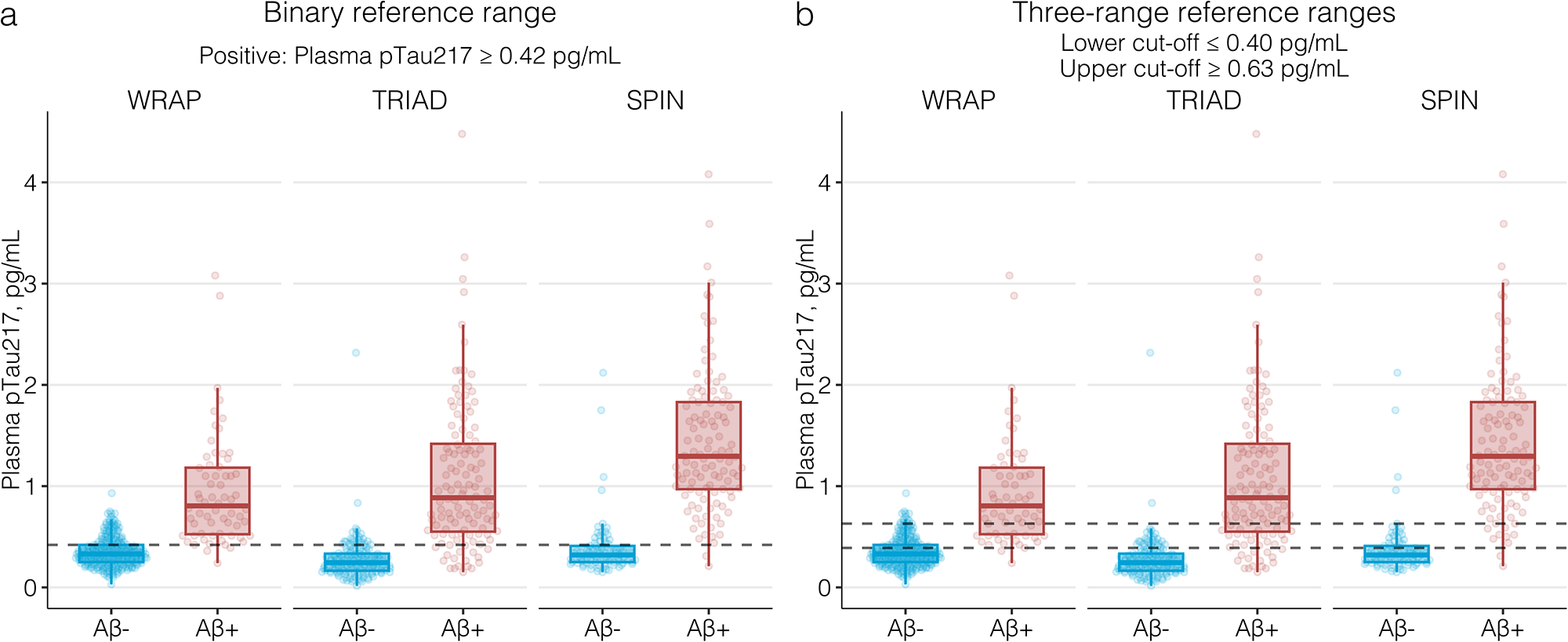
Binary and three-range ALZpath pTau217 reference ranges for Aβ-positivity. Boxplots show the distribution of plasma ALZpath pTau217 according to Aβ status (Aβ-negative: blue; Aβ-positive: red) for each of the three cohorts alongside reference ranges. (A) The dashed line represents a binary cut-off (>0.42 pg/mL) for Aβ-positivity derived based on the conventionally used Youden index. (B) The upper dashed line represents an upper cut-point (>0.63 pg/mL) for considering a pTau217 reading as positive, derived with 95% specificity for Aβ-positivity. The lower dashed line represents a lower cut-point (<0.40 pg/mL), below which plasma pTau217 would be considered negative, derived with 95% sensitivity for Aβ-positivity.

### Longitudinal change of ALZpath plasma pTau217

In up to 8 years of longitudinal sampling in WRAP (mean [SD] of 5.22 (1.41) years), the A+T+ group demonstrated a significantly higher annual increase rate in plasma pTau217 levels compared to the A-T-group (β-estimate: 0.12, 95%CI 0.10-0.13, *P*<0.0001). The A+T-group also demonstrated a significantly higher annual rate of change in plasma pTau217 compared to A-T-(β-estimate: 0.04, 95%CI 0.02-0.05, *P*<0.0001). Slope comparisons showed the A+T+ group to have a significantly higher rate compared to the A+T-group (β-estimate: 0.08; *P*<0.0001, **Figure 5A**). In TRIAD, similar results were observed, even with a shorter follow-up (1.90 [0.61] years). In comparison to the A-T-group, both A+T-(β-estimate: 0.07, 95%CI 0.03-0.11, *P*<0.001) and A+T+ (β-estimate: 0.13, 95%CI 0.07-0.19, *P*<0.0001) groups demonstrated significantly higher rates of change in plasma pTau217 levels. However, in contrast to WRAP, pairwise slope comparisons did not show significant differences between pTau217 rates of change between the A+T+ and A+T-groups (β-estimate: 0.06; *P*=0.20, **Figure 5B**).

**Figure 5.**
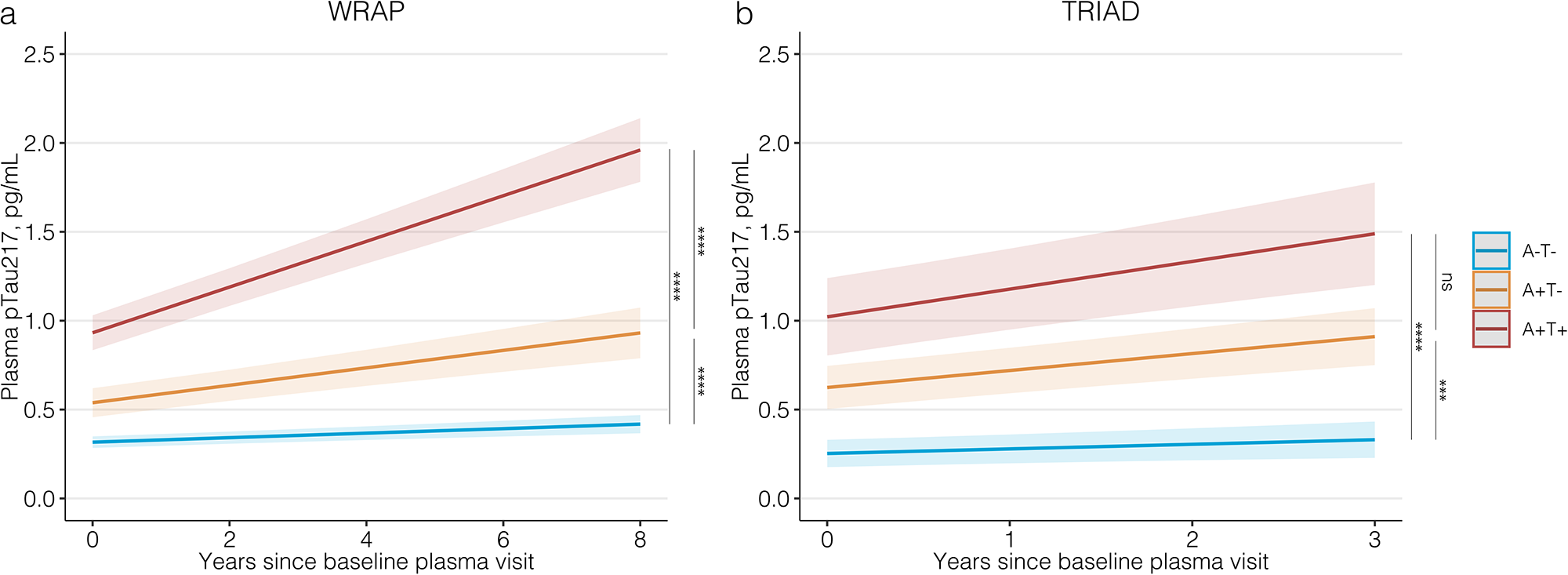
Longitudinal trajectories of plasma ALZpath pTau217 according to amyloid and tau PET status. Trajectory plots indicate the mean longitudinal trajectories (solid line) of plasma ALZpath pTau217 and associated 95% confidence intervals (ribbon), estimated with linear mixed-effects models. The left panel shows trajectories for the WRAP cohort (A) and the right panel for the TRIAD cohort (B). Trajectories are stratified based on PET-defined amyloid and tau (A-T-, A+T-, A+T+) groups, modeled with an interaction term between AT status and time. Models included random slopes and intercepts for each participant and were adjusted for years of education, sex, and cognitive status at first visit. P-values represent post-hoc pairwise comparing the slopes for group-by-time interactions. P<0.0001: ****; P<0.001: ***; P>0.05: not significant (ns).

## Discussion

In three independent cohorts, this study presents the clinical performance of the first widely available plasma pTau217 assay. Our findings demonstrate high accuracy in identifying abnormal Aβ and tau pathologies, comparable to CSF measures and superior to brain atrophy assessments. Notably, ALZpath pTau217 significantly outperforms other putative plasma biomarkers and their optimal combinations. Moreover, we found pTau217 cut-points for Aβ-positivity to be translatable across cohorts, being particularly effective in excluding the presence of Aβ pathology. A three-range approach demonstrated excellent negative and positive concordance with Aβ status, with approximately 20% of individuals falling into an “intermediate zone” that would require confirmatory testing via CSF or PET, as previously proposed^39^. Longitudinally, ALZpath pTau217 exhibited increases solely in individuals with Aβ pathology at baseline, and those with both Aβ and tau pathologies demonstrated a greater rate of increase.

Plasma biomarkers have emerged as crucial tools for AD evaluation. Their specificity to underlying pathology offers great potential for rapid screening, reducing the dependence on Aβ-PET scans or lumbar punctures in initial assessments. A clinical AD diagnosis often lacks sensitivity and specificity, resulting in many individuals with MCI (40–60%) or dementia (20–30%) exhibiting typical AD symptoms lacking brain Aβ pathology^1^. In primary care, where most patients with cognitive symptoms are initially managed, it is estimated that >50% of cognitive impaired patients remain undiagnosed or incorrectly diagnosed due to the lack of accessible and cost-effective tools^1^. Thus, blood biomarkers are set to revolutionise clinical care providing objective biomarker-based diagnostic information. This is especially important as anti-Aβ trials move toward targeting the preclinical population with lower prevalence of Aβ abnormalities^40^, in which a cost-effective screening strategy is paramount. In most previous studies, targeting pTau217 in blood has yielded the best results. Plasma pTau217 exhibits larger fold-changes in individuals with AD pathology, reducing uncertainty in three-range cut-off strategies, as explored here. Furthermore, pTau217 reflects both Aβ and tau pathologies and its longitudinal trajectory associates with cognitive decline and atrophy Aβ-positive individuals^24^.

A significant limitation of pTau217 has been the limited availability of immunoassays for broader evaluation. However, this study introduces a widely available plasma pTau217 assay exhibiting similar advantageous features to those observed in previous pTau217 studies. Consistent with Palmqvist et al^19^, ALZpath plasma pTau217 outperformed MRI features in identifying AD pathology, and showed comparable performance to CSF biomarkers in detecting Aβ-PET-positivity and tau-PET-positivity^41^. Impressively, it showed significant superiority to other validated plasma biomarkers and their optimal combinations. When combined with *APOE* status and age, the assay exhibited only modest improvements in diagnostic accuracy, whereas other biomarkers relied more heavily on these variables for their performance. Notably, the assay demonstrated high accuracy in identifying tau pathology within a larger group of Aβ-positive individuals. This is particularly important as anti-amyloid therapies may be less effective in patients with advanced tau pathology^42^, an inclusion strategy for the TRAILBLAZER trial in early AD, where Aβ-positive participants had to meet specific ranges of tau-PET burden^43^. Our findings suggest that pTau217 has the potential to identify elevated tau-PET uptake, making it a promising screening tool for early AD trials, and warranting further studies applying pTau217 to the “intermediate tau” designs such as that of the TRAILBLAZER trial.

Integrating blood biomarkers into diagnostic workflows remains challenging despite their excellent performance in identifying Aβ-positive patients. Therefore, this study also aimed to establish reference-points for ALZpath pTau217 based on abnormal Aβ pathology. The study evaluated a three-range approach, as recommended by Alzheimer’s Association guidelines^44^ and recently proposed by Brum et al using a non-commercial pTau217 immunoassay^39^, which suggests confirmatory testing (e.g., CSF or PET imaging) for patients with uncertain plasma results. Evaluating this approach using the ALZpath immunoassay showed high negative and positive predictive accuracy at screening, indicating only 12-23% would require further testing, depending on the clinical stage. However, we acknowledge that the cohorts utilized in this study may not fully represent real-world clinical settings. Importantly, the reported negative and positive predictive accuracy of these reference ranges can vary based on the prevalence of the outcome in the target population. Lower PPA’s are expected in settings with lower prevalence ^45^, as observed in the preclinical WRAP cohort compared to the higher prevalence seen in TRIAD and SPIN cohorts. Therefore, future studies should prospectively evaluate plasma pTau217 reference-points in memory clinic populations with wider diversity to ensure optimized implementation, accounting for higher rates of important co-morbidities^46^.

In conclusion, this study highlights the effectiveness of the commercially available plasma pTau217 assay in accurately identifying AD pathology. Our findings indicate that this immunoassay can significantly reduce the need for confirmatory testing for Aβ-positivity by approximately 80%, depending on their stage in the AD continuum. These results emphasize the crucial role of plasma pTau217 as an initial screening tool in the near future of AD management, and in the initiation and monitoring of anti-amyloid immunotherapies.

## Data Availability

All data produced in the present study are available upon reasonable request to the authors

## Acknowledgements

KB is supported by the Swedish Research Council (#2017-00915 and #2022-00732), the Swedish Alzheimer Foundation (#AF-930351, #AF-939721 and #AF-968270), Hjärnfonden, Sweden (#FO2017-0243 and #ALZ2022-0006), the Swedish state under the agreement between the Swedish government and the County Councils, the ALF-agreement (#ALFGBG-715986 and #ALFGBG-965240), the European Union Joint Program for Neurodegenerative Disorders (JPND2019-466-236), the Alzheimer’s Association 2021 Zenith Award (ZEN-21-848495), and the Alzheimer’s Association 2022-2025 Grant (SG-23-1038904 QC). HZ is a Wallenberg Scholar supported by grants from the Swedish Research Council (#2022-01018 and #2019-02397), the European Union’s Horizon Europe research and innovation programme under grant agreement No 101053962, Swedish State Support for Clinical Research (#ALFGBG-71320), the Alzheimer Drug Discovery Foundation (ADDF), USA (#201809-2016862), the AD Strategic Fund and the Alzheimer’s Association (#ADSF-21-831376-C, #ADSF-21-831381-C, and #ADSF-21-831377-C), the Bluefield Project, the Olav Thon Foundation, the Erling-Persson Family Foundation, Stiftelsen för Gamla Tjänarinnor, Hjärnfonden, Sweden (#FO2022-0270), the European Union’s Horizon 2020 research and innovation programme under the Marie Skłodowska-Curie grant agreement No 860197 (MIRIADE), the European Union Joint Programme – Neurodegenerative Disease Research (JPND2021-00694), the National Institute for Health and Care Research University College London Hospitals Biomedical Research Centre, and the UK Dementia Research Institute at UCL (UKDRI-1003). The Wisconsin Registry for Alzheimer’s Prevention is supported by NIH grants AG027161 and AG021155. JF, DA and AL were supported by the Fondo de Investigaciones Sanitario, Carlos III Health Institute (PI20/01473 to JF., PI18/00435 and INT19/00016 to DA, PI20/01330 to AL) and the Centro de Investigación Biomédica en Red sobre Enfermedades Neurodegenerativas (CIBERNED) Program 1. This work was also supported by the National Institutes of Health grants (1R01AG056850-01A1; R21AG056974 and R01AG061566 to JF) the Department de Salut de la Generalitat de Catalunya, Pla Estratègic de Recerca I Innovació en Salut (SLT006/17/00119 to JF; SLT002/16/00408 to AL; SLT006/17/125 to DA). It was also supported by Horizon 2020–Research and Innovation Framework Programme from the European Union (H2020-SC1-BHC-2018-2020 to JF). We thank Jeroen Vanbrabant and Erik Stoops from ADx Neuroscience, now Fujirebio, and the accelerator lab at Quanterix, for the initial prototyping of the ALZpath pTau217 assay.

