## Supplementary methods, Figures and Tables for "Diagnostic accuracy of the plasma ALZpath pTau217 immunoassay to identify Alzheimer’s disease pathology"

^1^ Department of Psychiatry and Neurochemistry, Institute of Neuroscience & Physiology, the Sahlgrenska Academy at the University of Gothenburg, Mölndal, Sweden; ^2^ King's College London, Institute of Psychiatry, Psychology and Neuroscience Maurice Wohl Institute Clinical Neuroscience Institute London UK; ^3^ NIHR Biomedical Research Centre for Mental Health and Biomedical Research Unit for Dementia at South London and Maudsley NHS Foundation London UK; ^4^ Centre for Age-Related Medicine, Stavanger University Hospital, Stavanger, Norway; ^5^ Graduate Program in Biological Sciences: Biochemistry, Universidade Federal do Rio Grande do Sul (UFRGS), Porto Alegre, Brazil; ^6^ Wisconsin Alzheimer's Institute, School of Medicine and Public Health, University of Wisconsin-Madison, Madison, WI 53726, USA; ^7^ Wisconsin Alzheimer's Disease Research Center, School of Medicine and Public Health, University of Wisconsin-Madison, Madison, WI 53792, USA; ^8^ Department of Medicine, Division of Geriatrics and Gerontology, School of Medicine and Public Health, University of Wisconsin-Madison, Madison, WI 53792, USA; ^9^ Geriatric Research Education and Clinical Center of the Wm. S. Middleton Memorial Veterans Hospital, Madison, WI 53705, USA; ^10^ ADx NeuroSciences, Technologiepark 94, Ghent, Belgium; ^11^ Translational Neuroimaging Laboratory, McGill University Research Centre for Studies in Aging, Alzheimer's Disease Research Unit, Douglas Research Institute, Le Centre intégré universitaire de santé et de services sociaux (CIUSSS) de l'Ouest-de-l'Île-de-Montréal; ^12^ Department of Neurology and Neurosurgery, Psychiatry and Pharmacology and Therapeutics, McGill University, Montreal, Quebec, Canada; ^13^ Department of Psychiatry, University of Pittsburgh, Pittsburgh, Pennsylvania, USA; ^14^ Department of Neurology, University of Pittsburgh, Pittsburgh, Pennsylvania, USA.; ^15^ Department of Neurology, Institut d'Investigacions Biomèdiques Sant Pau - Hospital de Sant Pau, Universitat Autònoma de Barcelona, Hospital de la Santa Creu i Sant Pau, Barcelona, Catalunya; ^16^ Centro de Investigación Biomédica en Red en Enfermedades Neurodegenerativas, CIBERNED, Madrid, Spain; ^17^ ALZpath. Inc, Carlsbad, CA 92008, USA; ^18^ Clinical Neurochemistry Laboratory, Sahlgrenska University Hospital, Mölndal, Sweden; ^19^ Department of Neurodegenerative Disease, UCL Institute of Neurology, London, UK; ^20^ UK Dementia Research Institute at UCL, London, UK; ^21^ Hong Kong Center for Neurodegenerative Diseases, Clear Water Bay, Hong Kong, China.

^†^ Nicholas J. Ashton and Wagner S. Brum contribute equally as 1^st^ authors

**Corresponding author**

Dr. Nicholas J. Ashton, Ph.D.

Institute of neuroscience and physiology

Dept. Psychiatry and Neurochemistry

Sahlgrenska Academy at Gothenburg University

Mölndal Hospital, Hus V3, 43180 Mölndal, Sweden

**Contents**

**Supplementary Methods**

The Translational Biomarkers in Aging and Dementia (TRIAD), p 4-5.

Wisconsin Registry for Alzheimer’s Prevention (WRAP), p 5-6.

Sant Pau Initiative on Neurodegeneration (SPIN), p 6-7.

**Supplementary Figures**

*Supplementary Figure 1* – Plasma ALZpath pTau217 levels according to amyloid status. p 8

*Supplementary Figure 2* – Plasma ALZpath pTau217 levels according to amyloid status defined based on PET visual reads. p 9.

*Supplementary Figure 3* **–** Plasma ALZpath pTau217 levels according to Braak stages in TRIAD., p 10.

*Supplementary Figure 4* –, Plasma ALZpath pTau217 also demonstrates similar or superior diagnostic accuracy for Aβ and tau pathologies compared to established CSF and PET biomarkers when evaluated in full biomarker availability subset. p 11

Supplementary Figure 5 –

Plasma pTau217 accuracy for discriminating tau pathology status among amyloid-positive individuals is superior to that of established biomarkers. p 12

Supplementary Figure 6 – Plasma pTau217 demonstrates higher accuracy for Aβ and tau positivity compared to other plasma biomarkers and their combinations. p 13

Supplementary Figure 7 – Correlations of ALZpath pTau217 with Aβ and tau PET. p 14

Supplementary Figure 8 –

Correlations of ALZpath pTau217 with CSF pTau217. p 15

**Supplementary Tables**

*Supplementary Table 1* – Demographics of longitudinal cohort. p 16

*Supplementary Table 2* – Intermediate precision and repeatability of the ALZpath pTau217 assay. p 17

*Supplementary Table 3* – ALZpath pTau217 levels by Braak stage. p 18

*Supplementary Table 4* – Receiver operating characteristics curves of plasma biomarkers to determine Aβ positivity. p 18

*Supplementary Table 5* – Receiver operating characteristics curves of plasma biomarkers to determine tau positivity. p 19

*Supplementary Table 6* – Binary reference values for for Aβ-positivity. p 21

*Supplementary Table 7* – Three-range reference values for Aβ-positivity. p 22

*Supplementary Table 9* – Binary reference values for Tau-positivity, pg 23

**Supplementary Methods**

*The Translational Biomarkers in Aging and Dementia (TRIAD)*

TRAID is an observational and longitudinal biomarker study approved by the Montreal Neurological Institute PET working committee and the Douglas Mental Health University Institute Research Ethics Board. Written informed consent was obtained for all participants. TRIAD participants are followed yearly with detailed clinical and neuropsychological assessments, as well as with collection of biofluids (blood, urine, saliva, and CSF) and acquisition of multiple imaging biomarkers. This study included cross-sectional data on 268 participants from TRIAD with multimodal imaging Aβ PET [^18^F]-AZD4694, tau PET [^18^F]-MK6240 and magnetic resonance imaging (MRI). A subset of participants with imaging had corresponding CSF biomarkers (Aβ42/40, pTau181, pTau205, ptau217, pTau231). The included participants were classified as cognitively unimpaired (CU, *n(%)* = 134(50%)), mild cognitive impairment (MCI, *n(%)* = 63(23.5%)), AD (*n(%)* = 46(17.2%)) and non-AD dementia (*n(%)* = 24(9.0%)). CU individuals had no objective cognitive impairment and a Clinical Dementia Rating (CDR) score of 0. Individuals with MCI had subjective and/or objective cognitive impairment and a CDR score of 0.5. Individuals with dementia had a CDR score of 1 or 2. Structural MRI data was acquired on a 3T Siemens Magnetom to obtain a high-resolution T1-weighted image of the entire brain. T1-weighted anatomical images were segmented using the SPM12 segmentation tool and non-linearly registered to the ADNI template using DARTEL, as previously reported ^1^. Brain atrophy was estimated using hippocampal volume, which measurements were estimated using FreeSurfer and were adjusted for total intracranial volume (ICV), as previously described ^2^. ICV adjustment was performed based on data from CU participants at baseline. T1-weighted anatomical images were also employed for coregistration purposes to PET images. A Siemens High Resolution Research Tomograph (HRRT) was used for PET imaging acquisitions, which occurred ±80 days from the CSF collection date (median = 53vdays). For Aβ PET, images were acquired 40–70 minutes post-injection of [^18^F]-AZD4694 and scans were reconstructed using the ordered subset expectation maximization (OSEM) algorithm on a 4-dimensional volume with 3 frames (3x600s) ^3^. For tau PET, [^18^F]-MK6240 scans were acquired 90–110 minutes post-injection and the OSEM algorithm was also used for reconstruction on a 4D volume with 4 frames (4x300s). Additional pre-processing corrections were performed as described elsewhere ^4^. PET images were meninges and skull stripped, linearly and non-linearly registered to the ADNI template space and then spatially smoothed to achieve a final resolution of 8 mm FWHM ^5^. The inferior cerebellum and whole cerebellum gray matter were used as the reference regions for [^18^F]-MK6240 and [^18^F]-AZD4694, respectively. Global Aβ PET used averaged SUVR of the precuneus, cingulate, inferior parietal, medial prefrontal, lateral temporal, and orbitofrontal cortices and a positivity value of 1.55 ^6^, corresponding to 24 Centiloids ^7^. Aβ positivity was also visually defined by two neurologists blinded to clinical diagnosis. Tau PET SUVR was globally estimated from a composite area including the meta-ROI region. Tau positivity was defined 1.24 as previously described ^8^ (mean + 2 standard deviations (SD) higher than the mean meta-ROI region of the young (<26 years of age) participants).

CSF samples were collected by syringe and transferred to polypropylene tubes for centrifugation at 20 °C, 2200g for 10 minutes. Samples were then distributed into 1 millilitre aliquots in polypropylene vials (Fisher Scientific Inc. Catalog # 3741-WP1D-BR) and permanently stored at -80 °C pending biochemical analyses at the Department of Neurochemistry, University of Gothenburg. CSF pTau181 and Aβ42/40 were quantified by the LUMIPULSE G1200 as previously described ^6^. CSF pTau217 ^9^ and pTau205 (Lantero-Rodriguez et al., unpublished) were quantified by in-house Simoa assay developed at the University of Gothenburg. All plasma analysed for TRIAD was performed at the Department of Psychiatry and Neurochemistry, University of Gothenburg. Plasma Aβ42/40, GFAP and NfL were quantified by the commercial Neurology 4-plex E (#103670, Quanterix). Plasma pTau181 and pTau231 were analysed by in-house Simoa assays developed at the University of Gothenburg ^10, 11^. Plasma pTau217 was quantified by the ALZpath Simoa assay as described in the manuscript methods at the Department of Psychiatry and Neurochemistry, University of Gothenburg between 1^st^ December 2022 and 22^nd^ December 2022.

*Wisconsin Registry for Alzheimer’s Prevention (WRAP)*

The WRAP study and the measures collected for this project were approved by the University of Wisconsin-Madison institutional review board and written informed consent was obtained for all participants. WRAP ^12^ is an observational longitudinal observational cohort that collects biofluid, cognitive and clinical data at approximate biennial visits. An expanding subset of participants undergo amyloid PET [^11^C]-PiB, tau PET [^18^F]-MK6240, MRI and/or CSF collection procedures. Participants who had provided suitable plasma (with EDTA anticoagulant which began in 2011) and had undergone at least one lumbar puncture or at least one [^11^C]-PiB were eligible to be included. The majority of those with [^11^C]-PiB data also underwent tau PET with [^18^F]-MK6240. This study included cross-sectional data on 323 WRAP participants, and mostly CU (CU, *n(%)* = 309(95.6%) at their first available plasma sample collection with no clinically significant cognitive impairment based on a consensus panel review. Some individuals at first available plasma sample draw had cognitive impairment MCI, *n(%)* = 12(3.7%); dementia, *n(%)* = 2(0.6%)) using standard diagnostic criteria. All participants with PET also had acquired 3T T1-weighted images for co-registration purposes. PET imaging was acquired on a Siemens HR+. For Aβ PET, images were acquired dynamically 0–70 minutes post-injection of PiB as described previously from which mean cortical distribution volume ratios (DVR) was derived using Logan graphical analysis with the cerebellum gray matter as the reference region ^13^. For tau PET, [^18^F]-MK6240 scans were acquired 70-90 minutes post-injection and meta-temporal ROI standard uptake value ratios (SUVR) were derived as described elsewhere ^14, 15^. PET images were spatially registered with the MNI template and the AAL3 (in the case of PiB) or Harvard Oxford atlas (for tau PET) standard regions were extracted. For WRAP the global cortical Aβ PET utilized the averaged DVR of the precuneus, anterior and posterior cingulate, inferior parietal, medial prefrontal, lateral temporal, and orbitofrontal cortices and positivity was defined as a DVR of 1.19 or greater which corresponded to a Centiloid >21.7) as described elsewhere ^16^. For the purposes of this study, an additional Aβ PET Centiloid of 24 was used for purposes of comparison to the TRIAD cohort. Tau PET SUVR was assessed in the temporal meta-ROI, a commonly used summary measure of tau PET ^17^ that encompasses the entorhinal cortex, amygdala, parahippocampal gyrus, fusiform gyrus, inferior and middle temporal gyrus. Tau positivity was defined as [^18^F]-MK6240 temporal meta-ROI SUVR > 1.30 (unpublished) which was 2.5 standard deviations (SD) higher than the mean of middle-aged (< 60), amyloid negative participants.

CSF samples were collected by syringe and transferred to polypropylene tubes for centrifugation at 20 °C, 2200 g for 10 min. Samples were then distributed into 0.5mL aliquots in polypropylene vials and permanently stored at -80 °C pending analyses. CSF biomarkers (Aβ42, Aβ40, pTau181 among others) using the Roche NeuroToolKit ^18^ were measured at the Department of Psychiatry and Neurochemistry, University of Gothenburg. All plasma analysed for this study was performed at the Department of Psychiatry and Neurochemistry, University of Gothenburg, and identical to TRIAD, with exception of plasma pTau181, which was quantified by the commercial pTau-181 Advantage V2.1 Simoa (#104111, Quanterix). Plasma pTau217 was quantified by the ALZpath Simoa assay as described in the manuscript methods at the Department of Psychiatry and Neurochemistry, University of Gothenburg between 5^th^ Janurary 2023 and 3^rd^ February 2023.

*Sant Pau Initiative on Neurodegeneration (SPIN)*

The SPIN cohort is a comprehensive observational platform for studying neurodegenerative diseases that uses multiple types of biomarkers and takes an integrative approach ^19^. Individuals who participate in SPIN agree to donate biofluid (blood and CSF) and undergo detailed neurological and neuropsychological evaluations. A subset of participants also receives a 3T brain MRI scan and additional functional or imaging studies such as a video-polysomnogram, [^18^F]-fluorodeoxyglucose PET, amyloid PET, or Tau PET. Participants are followed for at least 4 years, with additional samples and imaging studies taken every other year. All procedures in the study were approved by the ethics committee of Hospital Sant Pau, and all participants or their legally authorized representatives provided written informed consent in accordance with the Declaration of Helsinki. For cross-sectional analysis, we included 195 participants with biomarker confirmation (CSF or PET). We included individuals with mild cognitive impairment due to (MCI-AD, *n(%)* = 72(36.7%)), AD dementia (AD, *n(%)* = 41(21.0%)) and and cognitively unimpaired controls (CU, *n(%)* = 82(42.1%)). Diagnosis was based on internationally recognized clinical criteria, and control participants had normal cognitive scores on standard neuropsychological evaluations. A subset of participants had structural 3T-MRI and/or amyloid PET ([^18^F]-florbetapir or [^18^F]-flutemetamol, n = 32). The Computational Anatomy Toolbox (CAT12, http://dbm.neuro.uni-jena.de/cat), which is a tool for the SPM12 software, was used to pre-process the structural T1 sequence of the MRI and to extract hippocampal volumes ^20^. PET images were co-registered to the corresponding MRI of everyone. The amyloid PET images were intensity-scaled using the cerebellum region. All resulting PET images were projected to the middle point of the cortical ribbon, inspected visually for potential errors, and smoothed with a 10mm kernel. We normalized the amyloid PET images to the MNI space and calculated the standardized uptake value ratio (SUVR) using the target amyloid region and whole cerebellum regions of the GAAIN website (www.gaain.org). The SUVR values were then converted to the Centiloid scale ^21^.

CSF samples were collected in 10 mL polypropylene tubes (Sarstedt, #62.610.018), which were then taken to the Sant Pau Memory Unit laboratory. Within 2 hours, samples were centrifuged, aliquoted, and stored at -80°C. CSF levels of core AD biomarkers (Aβ42, Aβ40, and pTau181) were measured in the Lumipulse fully-automated platform using commercially available kits (Fujirebio Europe, Ghent, Belgium), as previously described ^21^.

Blood samples were collected in 10 ml EDTA-2K tubes and then centrifuged for 10 minutes at 4°C. All plasma analysed for this study was performed at the Department of Psychiatry and Neurochemistry, University of Gothenburg, and identical to TRIAD. Plasma pTau217 was quantified by the ALZpath Simoa assay as described in the manuscript methods at the Department of Psychiatry and Neurochemistry, University of Gothenburg between 20^th^ March 2023 and 30^rd^ March 2023.

**Supplementary Figure 1**. Plasma ALZpath pTau217 levels according to amyloid status.

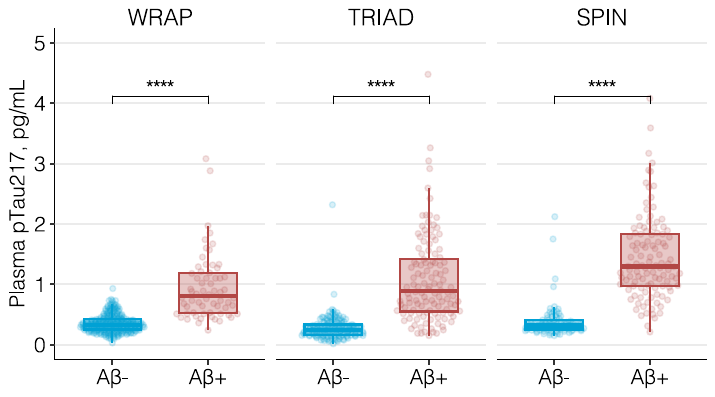

Boxplots show the distribution of ALZpath pTau217 value by Aβ profile for the WRAP, TRIAD, and SPIN cohorts. For WRAP and TRIAD, Aβ (“A”) was indexed by PET. In SPIN, Aβ (“A”) was indexed by CSF Aβ42/40. P-values were obtained from pairwise contrasts performed on linear models controlling for age and sex, and all were <0.0001 (****), with Tukey multiplicity adjustment.

**Supplementary Figure 2.** Plasma ALZpath pTau217 levels according to amyloid status defined based on PET visual reads.

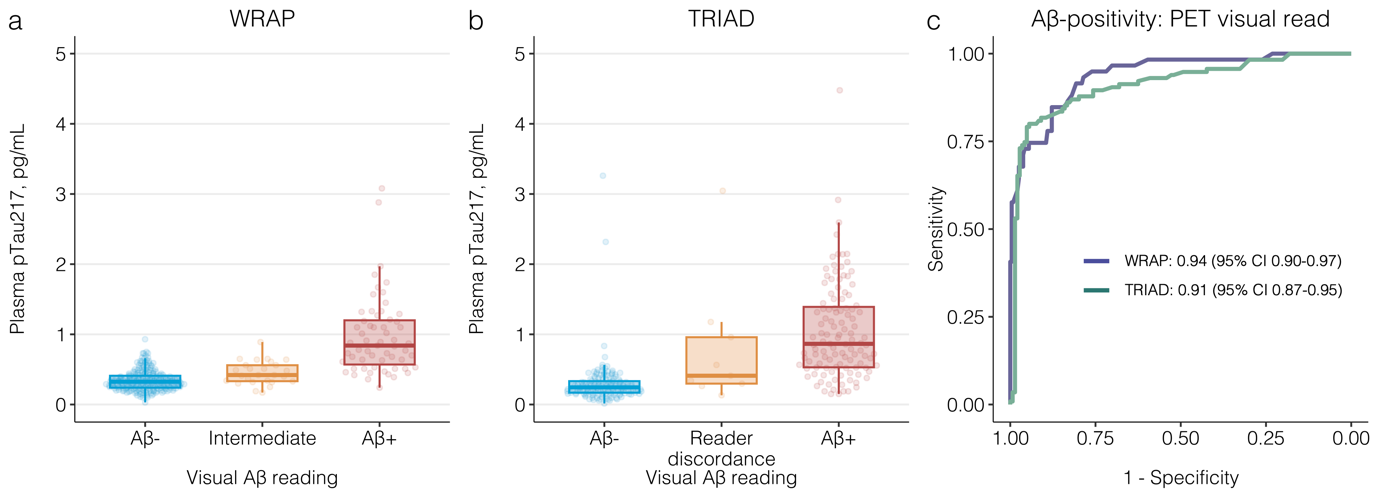

Boxplots show the distribution of ALZpath pTau217 value by visual Aβ PET reading for WRAP (A) and TRIAD (B). Receiver operating characteristics (ROC) curves for ALZpath pTau217 for Aβ positivity by visual read in both WRAP and TRIAD (C). For each ROC curve, the area under the curve (AUC) is reported alongside 95% confidence intervals (CI).

**Supplementary Figure 3 –** Plasma ALZpath pTau217 levels according to Braak stages in TRIAD.

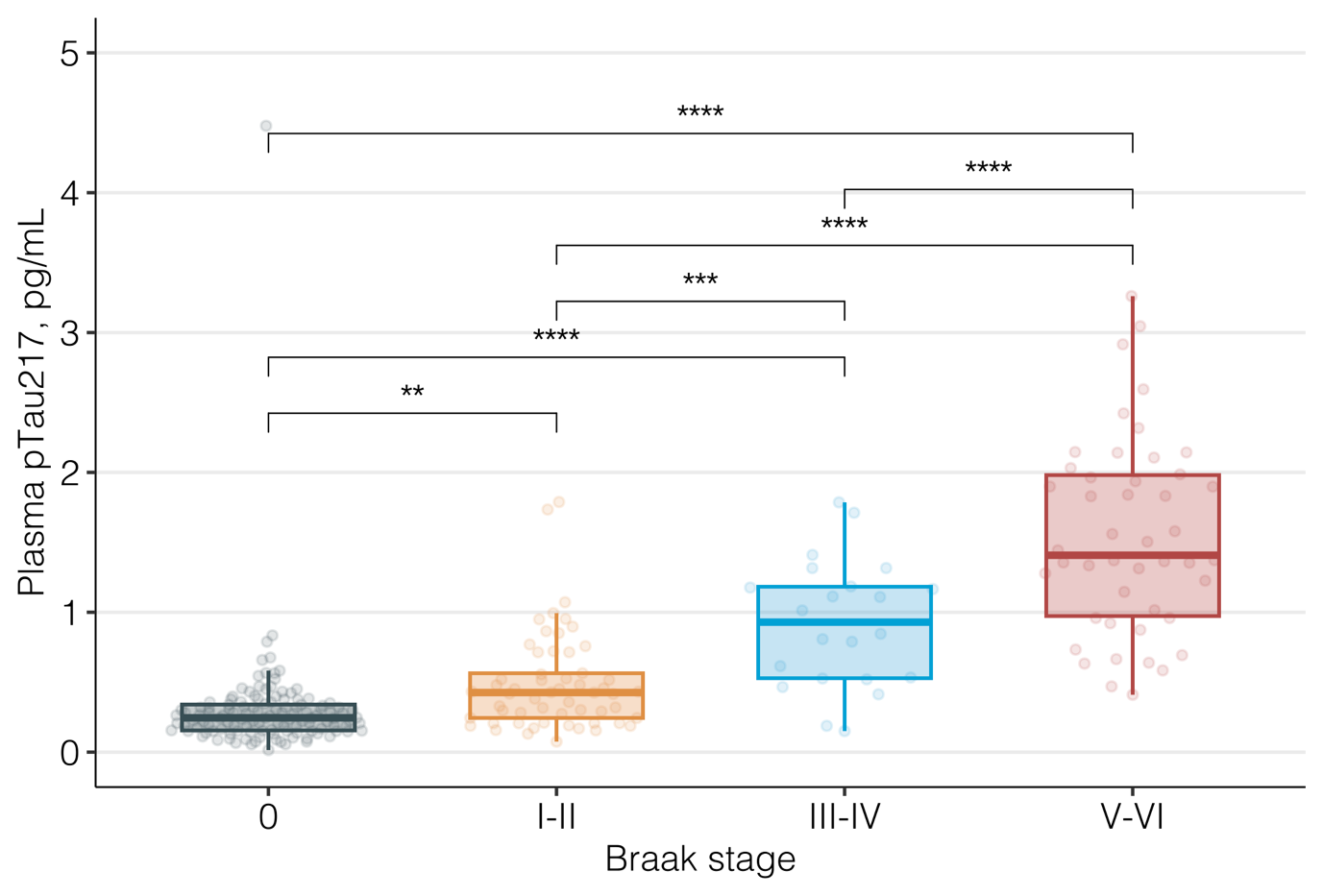

Boxplots show the distribution of ALZpath pTau217 levels by Braak stage determined by tau PET profile for the TRIAD cohort. P-values were obtained from pairwise contrasts performed on a linear model adjusted for age and sex, with Tukey multiplicity adjustment.

P _0vsI-II_=0.22 (ns); P _I-IIvsIII-IV_=0.0042 (***); all others P<0.0001 (****).

**Supplementary Figure 4.** Plasma ALZpath pTau217 also demonstrates similar or superior diagnostic accuracy for Aβ and tau pathologies compared to established CSF and PET biomarkers when evaluated in full biomarker availability subset.

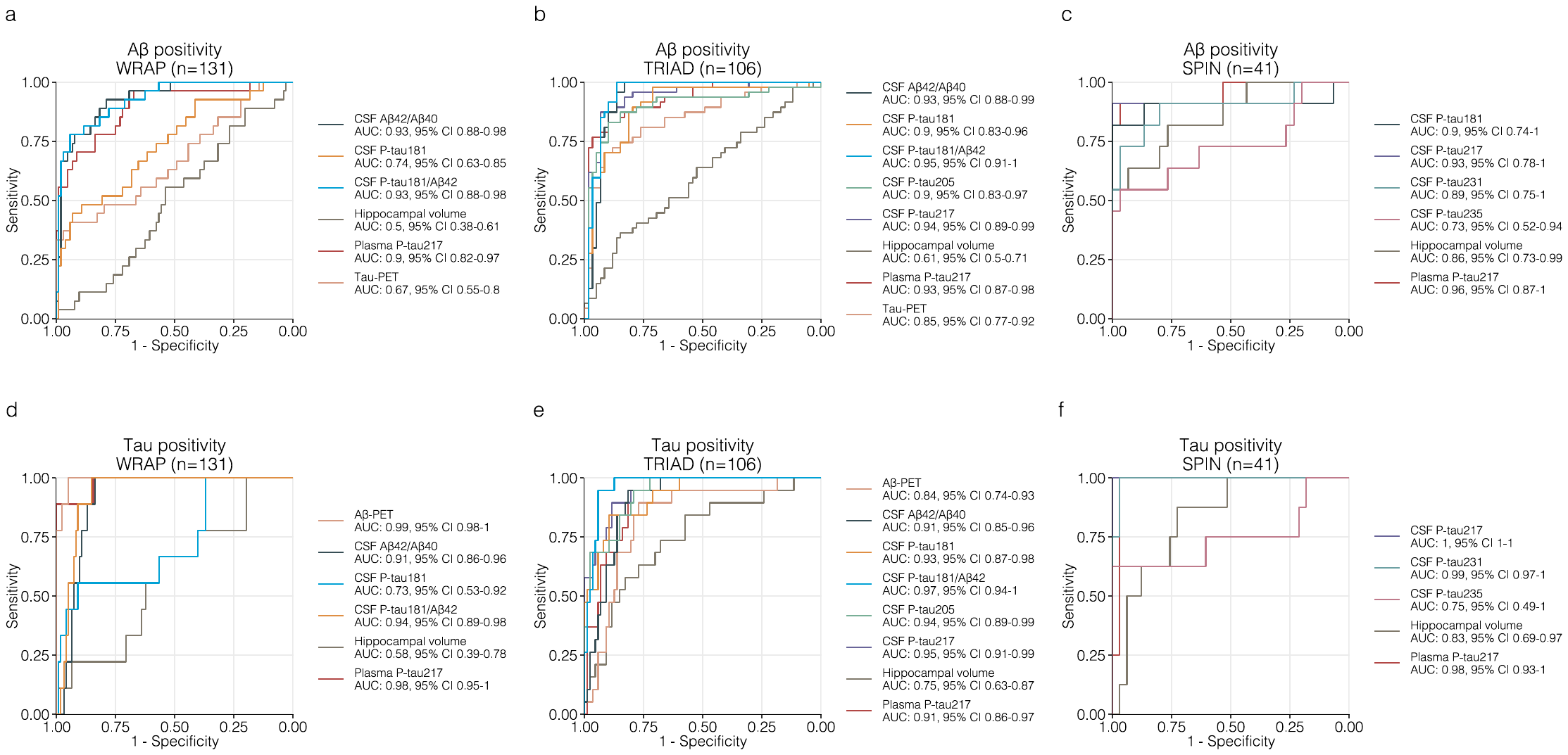

Receiver operating characteristics curves for detecting Aβ (A-C) and tau-positivity (D-F) for ALZpath pTau217, CSF and imaging biomarkers. Each panel corresponds to accuracies for the same outcome at each cohort (WRAP: A, D; TRIAD: B, E; SPIN: C, F). For WRAP and TRIAD, Aβ (“A”) and tau (“T”) were indexed by PET. In SPIN, “A” was indexed by CSF Aβ42/40 and “T” by pTau181. Solid lines represent the ROC curves for each biomarker, with colors corresponding to a specific biomarker across cohorts, as indicated in the figure legend at each panel. The legend indicates the area under the curve (AUC) for each biomarker, alongside 95% confidence intervals (CI). As indicated in the title of each panel, this analysis was conducted in a same subset of participants who had available data for all biomarkers included in the comparison. This approach led to similar results to the one presented in the main text, in which no data is excluded and the maximum number of observations per biomarker was used.

**Supplementary Figure 5.** Plasma pTau217 accuracy for discriminating tau pathology status among amyloid-positive individuals is superior to that of established biomarkers.

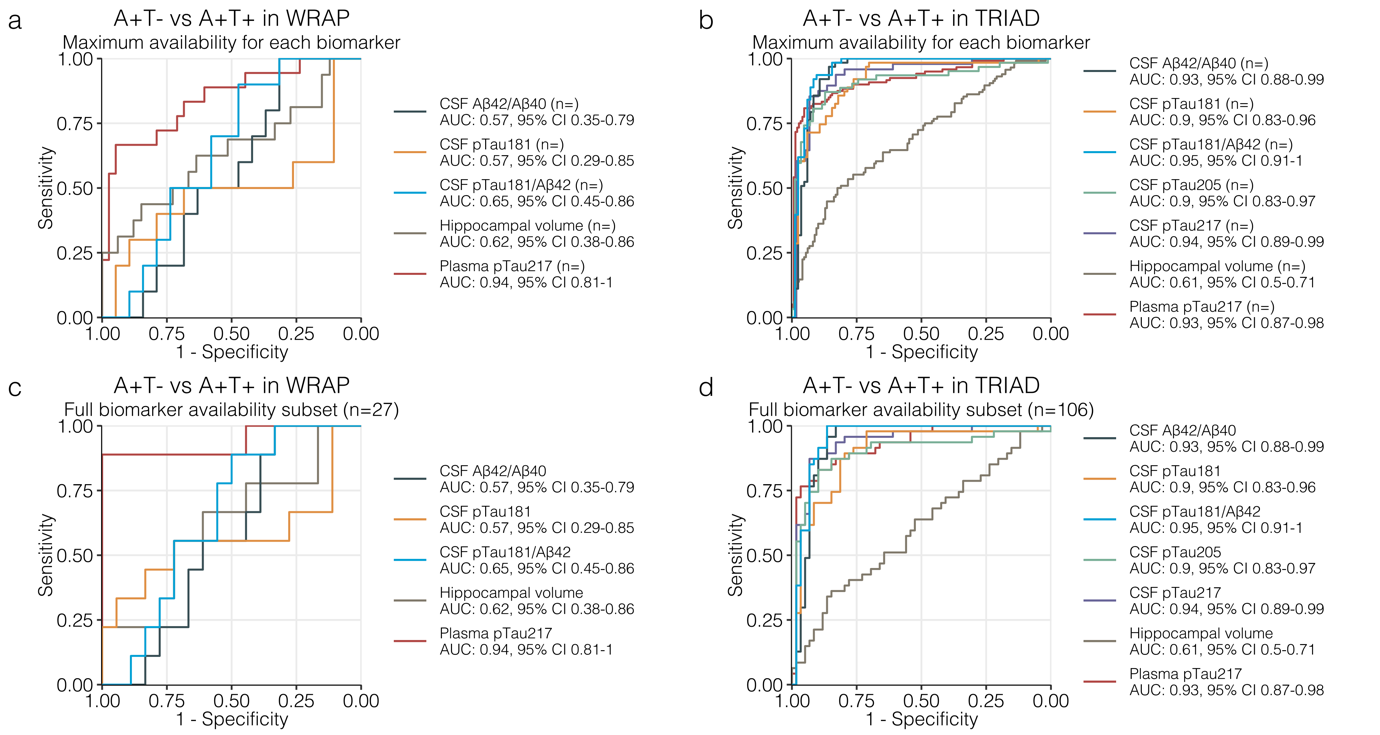

Receiver operating characteristics (ROC) curves for ALZpath pTau217 for discriminating A+T+ from A+T- in WRAP (A, C) and TRIAD (B, D), both modalities defined by PET. In (A) and (B), the analyses were performed based on the maximum number of available observations for each biomarker-outcome combination, while in (C) and (D) analyses were conducted in reduced subsets with those participants who had available data for all biomarkers. The legend indicates the area under the curve (AUC) for each biomarker, alongside 95% confidence intervals (CI).

**Supplementary Figure 6.** Plasma pTau217 demonstrates higher accuracy for Aβ and tau positivity compared to other plasma biomarkers and their combinations.

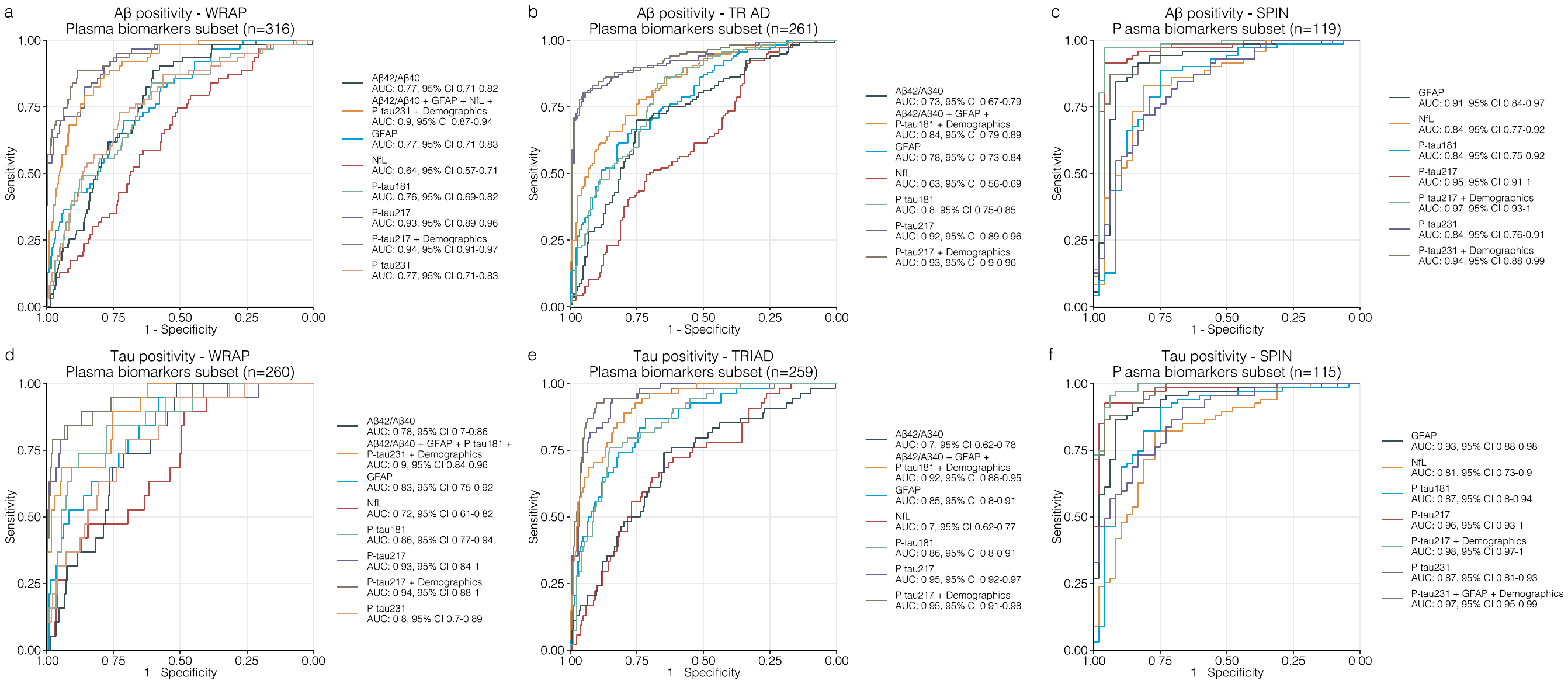

Receiver operating characteristics (ROC) curves for plasma biomarkers and plasma biomarker combinations to detect Aβ positivity (A-C) and tau positivity (D-F). The legend indicates the area under the curve (AUC) for each plasma biomarker or plasma biomarker combination, alongside 95% confidence intervals (CI). For WRAP and TRIAD, Aβ (“A”) and tau (“T”) were indexed by PET. In SPIN, “A” was indexed by CSF Aβ42/40 and “T” by pTau181. Biomarker combinations were evaluated based on model metrics shown in Supplementary Tables 4-5. “Demographics” refers to the addition of age, sex and *APOE* status.

**Supplementary Figure 7.** Correlations of ALZpath pTau217 with Aβ and tau PET.

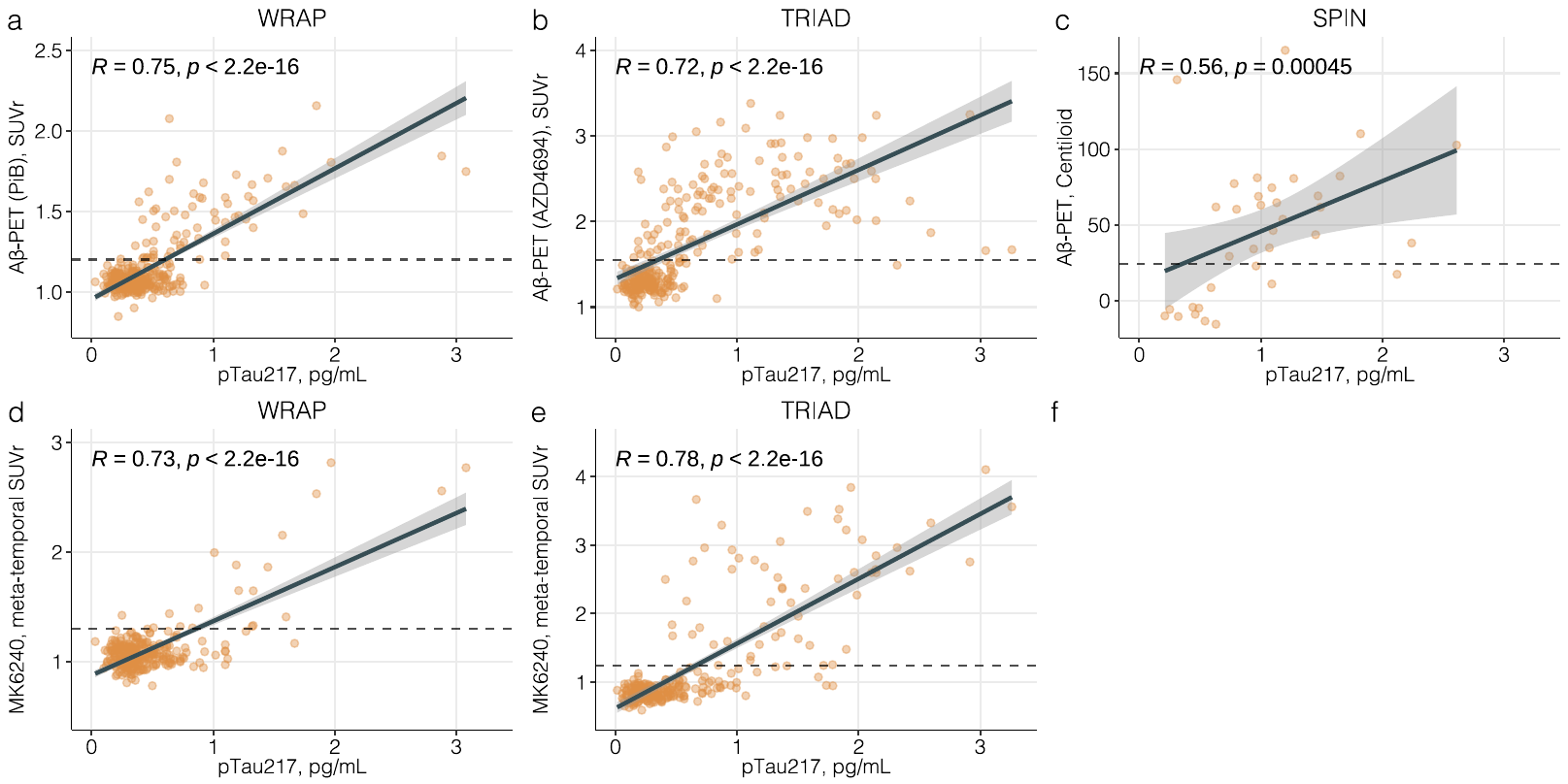

Correlations of ALZpath pTau217 with Aβ PET in WRAP (A), TRIAD (B) and SPIN (C), and between pTau217 and tau-PET in WRAP (D) and TRIAD (E). Correlation coefficients correspond to Spearman’s rho. The Aβ- or tau-PET ligand is indicated in the y-axis, except for SPIN, in which Aβ-PET is represented in the centiloid scale since patients underwent [^18^F]-flutemetamol or [^18^F]-florbetapir.

**Supplementary Figure 8.** Correlations of ALZpath pTau217 with CSF pTau217.

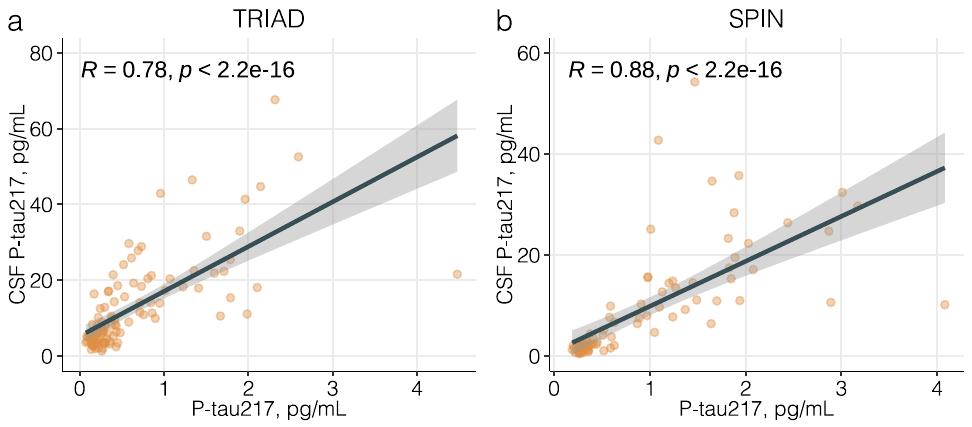

Correlations of ALZpath pTau217 with CSF pTau217, TRIAD (A) and SPIN (B). Correlation coefficients correspond to Spearman’s rho. CSF pTau217 measures were not available in WRAP.

**Supplementary Table 1** – Demographics of longitudinal cohort.

|  | WRAP | | | TRIAD | | |
| --- | --- | --- | --- | --- | --- | --- |
|  | A-T- (N=217) | A+T- (N=26) | A+T+ (N=17) | A-T- (N=80) | A+T- (N=40) | A+T+ (N=12) |
| Age, years, mean (SD) | 61.5 (6.54) | 64.0 (5.26) | 65.0 (5.71) | 70.5 (5.94) | 73.8 (5.04) | 70.1 (5.58) |
| Female, n (%) | 144 (66.4%) | 15 (57.7%) | 14 (82.4%) | 50 (62.5%) | 25 (62.5%) | 8 (66.7%) |
| APOE ε4 carriers, n (%) | 75 (34.6%) | 17 (65.4%) | 13 (76.5%) | 21 (26.3%) | 14 (35.0%) | 8 (66.7%) |
| Baseline MMSE score,  mean (SD) | 29.4 (0.887) | 29.0 (1.22) | 27.5 (3.10) | 29.0 (1.27) | 28.5 (1.39) | 27.3 (2.00) |
| Baseline clinical diagnosis |  |  |  |  |  |  |
| CU, n (%) | 212 (97.7%) | 26 (100%) | 17 (100%) | 70 (87.5%) | 21 (52.5%) | 0 (0%) |
| CI, n (%) | 5 (2.3%) | 0 (0%) | 0 (0%) | 10 (12.5%) | 19 (47.5%) | 12 (100%) |
| Years of education,  mean (SD) | 16.2 (2.76) | 16.8 (2.15) | 15.7 (2.23) | 15.5 (3.83) | 15.1 (3.16) | 14.7 (3.70) |
| Baseline plasma pTau217,  pg/mL, mean (SD) | 0.333 (0.142) | 0.553 (0.339) | 0.895 (0.395) | 0.260 (0.141) | 0.808 (0.744) | 1.07 (0.447) |
| Mean (SD) years of follow-up | 5.28 (1.38) | 5.06 (1.47) | 4.76 (1.71) | 1.95 (0.618) | 1.87 (0.620) | 1.68 (0.546) |
| Median (IQR) number of samples | 3 (3-4) | 3 (3-3) | 3 (2-3) | 2 (2-3) | 2 (2-3) | 2 (2-2.25) |

Data are mean (SD) or n (%). In both cohorts, AT status was defined with amyloid and tau-PET. Abbreviations: SD, standard deviation; TRIAD, Translational Biomarkers in Aging and Dementia; WRAP, Wisconsin Registry for Alzheimer’s Prevention; MMSE, mini-metal state examination; CU, cognitively unimpaired; CI, cognitively impaired; A-T-, amyloid-negative and tau-negative; AT-, amyloid-positive and tau-negative; A-T+, amyloid-positive and tau-positive.

**Supplementary Table 2** – Intermediate precision and repeatability of the ALZpath pTau217 assay.

| Quality Controls (range, pg/mL) | Repeatability (% CV_r_); Intermediate precision (%CV_Rw_) | | |
| --- | --- | --- | --- |
|  | WRAP | TRIAD | SPIN |
| IQC-1 (0.5–0.7) | 8.1; 8.2 | 10.6; 13.6 | 4.5; 8.1 |
| IQC-2 (1.6–2.0) | 6.9; 9.6 | 10.3; 17.9 | 4.0; 7.6 |
| IQC-3 (2.3) | 6.6; 9.7 | 8.2;13.5 | 5.3; 7.1 |
| EQC-1 (0.77–0.88) | 1.5; 8.8 | 1.1;3.6 | 1.7; 7.1 |
| EQC-2 (0.14- 0.2) | 3.5; 11.0 | 1.9;6.9 | 2.2; 7.5 |

Precision of plasma ALZpath pTau217 assay in all cohorts. IQC samples are human plasma samples from the University of Gothenburg. EQC samples are plasma samples provided by ALZpath commercial assay.

**Supplementary Table 3** – ALZpath pTau217 levels by Braak stage in the TRIAD cohort.

| Braak Stage | n | ALZpath pTau217, pg/mL (SD) |
| --- | --- | --- |
| 0 | 124 | 0.30 (0.41) |
| I-II | 57 | 0.49 (0.35) |
| III-IV | 22 | 0.92 (0.46) |
| V-VI | 46 | 1.55 (0.70) |

The table indicates the mean (SD) concentrations of plasma ALZpath pTau217 in the TRIAD cohort, According to Braak stages defined with [^18^F]MK-6240.

**Supplementary Table 4** – Receiver operating characteristics curves of plasma biomarker to determine Aβ-status.

| **Cohort** | **Biomarker** | **AUC** | **CI Lower** | **CI Upper** | **AIC** |
| --- | --- | --- | --- | --- | --- |
| WRAP | pTau217 | 0.931 | 0.897 | 0.965 | 161 |
|  | pTau231 | 0.774 | 0.711 | 0.838 | 274 |
|  | Aβ42/Aβ40 | 0.770 | 0.713 | 0.827 | 284 |
|  | GFAP | 0.766 | 0.704 | 0.828 | 271 |
|  | pTau181 | 0.765 | 0.701 | 0.829 | 289 |
|  | NfL | 0.650 | 0.581 | 0.720 | 312 |
|  | Demographics + pTau217 | 0.940 | 0.905 | 0.974 | 148 |
|  | Demographics + Aβ42/Aβ40 + GFAP + NfL + pTau231 | 0.903 | 0.866 | 0.940 | 209 |
|  | Demographics + Aβ42/Aβ40 + GFAP + NfL + pTau181+ pTau231 | 0.904 | 0.867 | 0.941 | 211 |
|  | Demographics + Aβ42/Aβ40 + GFAP + pTau231 | 0.893 | 0.853 | 0.933 | 215 |
|  | Demographics + Aβ42/Aβ40 + GFAP + pTau181+ pTau231 | 0.894 | 0.855 | 0.934 | 217 |
|  | Demographics + Aβ42/Aβ40 + GFAP + NfL + pTau181 | 0.895 | 0.859 | 0.932 | 219 |
| TRIAD | pTau217 | 0.923 | 0.888 | 0.957 | 193 |
|  | pTau181 | 0.798 | 0.745 | 0.851 | 301 |
|  | GFAP | 0.783 | 0.729 | 0.838 | 292 |
|  | Aβ42/Aβ40 | 0.736 | 0.676 | 0.797 | 324 |
|  | NfL | 0.624 | 0.557 | 0.692 | 355 |
|  | Demographics + pTau217 | 0.929 | 0.896 | 0.961 | 193 |
|  | Demographics + Aβ42/Aβ40 + GFAP + pTau181 | 0.861 | 0.817 | 0.904 | 253 |
|  | Demographics + Aβ42/Aβ40 + GFAP + NfL + pTau181 | 0.862 | 0.819 | 0.906 | 254 |
|  | Demographics + Aβ42/Aβ40 + GFAP | 0.840 | 0.793 | 0.888 | 265 |
|  | Demographics + Aβ42/Aβ40 + GFAP + NfL | 0.841 | 0.794 | 0.888 | 266 |
|  | Demographics + Aβ42/Aβ40 + pTau181 | 0.841 | 0.794 | 0.888 | 268 |
| SPIN | pTau217 | 0.957 | 0.924 | 0.990 | 69 |
|  | GFAP | 0.904 | 0.854 | 0.953 | 103 |
|  | NfL | 0.865 | 0.802 | 0.929 | 123 |
|  | pTau181 | 0.865 | 0.807 | 0.922 | 129 |
|  | pTau231 | 0.860 | 0.805 | 0.916 | 122 |
|  | Demographics + pTau217 | 0.969 | 0.931 | 1.000 | 59 |
|  | Demographics + GFAP + pTau231 | 0.940 | 0.887 | 0.994 | 82 |
|  | Demographics + pTau231 | 0.935 | 0.884 | 0.987 | 83 |
|  | Demographics + NfL + pTau231 | 0.940 | 0.887 | 0.993 | 83 |
|  | Demographics + GFAP | 0.938 | 0.887 | 0.988 | 84 |
|  | Demographics + GFAP + NfL + pTau231 | 0.939 | 0.884 | 0.993 | 84 |

The table demonstrated the area under the curve (AUC) for plasma biomarkers and their combinations, in logistic regression models, and their associated 95% confidence intervals (CI) for predicting Aβ-positivity (defined with Aβ-PET in WRAP and TRIAD, and with CSF Aβ42/Aβ40 in SPIN). The Akaike information criterion (AIC) is also shown.

**Supplementary Table 5** – Receiver operating characteristics curves of plasma biomarker to determine tau positivity.

| **Cohort** | **Biomarker** | **AUC** | **CI Lower** | **CI Upper** | **AIC** |
| --- | --- | --- | --- | --- | --- |
| WRAP | pTau217 | 0.927 | 0.845 | 1.000 | 71 |
|  | pTau181 | 0.855 | 0.767 | 0.944 | 118 |
|  | GFAP | 0.837 | 0.755 | 0.918 | 116 |
|  | pTau231 | 0.800 | 0.707 | 0.894 | 117 |
|  | Aβ42/Aβ40 | 0.776 | 0.696 | 0.856 | 126 |
|  | NfL | 0.719 | 0.611 | 0.826 | 133 |
|  | Demographics + Aβ42/Aβ40 + GFAP + pTau181+ pTau231 | 0.901 | 0.838 | 0.963 | 100 |
|  | Demographics + Aβ42/Aβ40 + GFAP + NfL + pTau181 | 0.903 | 0.842 | 0.964 | 101 |
|  | Demographics + Aβ42/Aβ40 + GFAP + NfL + pTau181+ pTau231 | 0.903 | 0.842 | 0.963 | 102 |
|  | Demographics + Aβ42/Aβ40 + pTau181+ pTau231 | 0.896 | 0.832 | 0.960 | 103 |
|  | Demographics + pTau217 | 0.941 | 0.880 | 1.000 | 71 |
|  | Demographics + Aβ42/Aβ40 + GFAP + pTau181 | 0.898 | 0.835 | 0.962 | 99 |
| TRIAD | pTau217 | 0.946 | 0.920 | 0.973 | 156 |
|  | pTau181 | 0.855 | 0.804 | 0.906 | 210 |
|  | GFAP | 0.851 | 0.796 | 0.905 | 200 |
|  | Aβ42/Aβ40 | 0.696 | 0.617 | 0.775 | 250 |
|  | NfL | 0.696 | 0.621 | 0.771 | 262 |
|  | Demographics + pTau217 | 0.947 | 0.911 | 0.982 | 152 |
|  | Demographics + Aβ42/Aβ40 + GFAP + pTau181 | 0.920 | 0.885 | 0.955 | 162 |
|  | Demographics + Aβ42/Aβ40 + GFAP + NfL + pTau181 | 0.920 | 0.884 | 0.955 | 164 |
|  | Demographics + GFAP + pTau181 | 0.906 | 0.865 | 0.946 | 170 |
|  | Demographics + GFAP + NfL + pTau181 | 0.906 | 0.865 | 0.946 | 172 |
|  | Demographics + Aβ42/Aβ40 + GFAP | 0.912 | 0.874 | 0.949 | 175 |
| SPIN | pTau217 | 0.970 | 0.944 | 0.997 | 56 |
|  | GFAP | 0.906 | 0.858 | 0.954 | 81 |
|  | pTau181 | 0.905 | 0.858 | 0.953 | 115 |
|  | pTau231 | 0.886 | 0.837 | 0.935 | 105 |
|  | NfL | 0.834 | 0.764 | 0.904 | 126 |
|  | Demographics + pTau217 | 0.984 | 0.966 | 1.000 | 47 |
|  | Demographics + GFAP + NfL + pTau231 | 0.974 | 0.952 | 0.996 | 62 |
|  | Demographics + GFAP + pTau231 | 0.970 | 0.946 | 0.994 | 63 |
|  | Demographics + GFAP + NfL + pTau181+ pTau231 | 0.973 | 0.951 | 0.995 | 64 |
|  | Demographics + GFAP + pTau181+ pTau231 | 0.970 | 0.946 | 0.994 | 65 |
|  | Demographics + pTau231 | 0.961 | 0.929 | 0.992 | 68 |

The table demonstrated the area under the curve (AUC) for plasma biomarkers and their combinations, in logistic regression models, and their associated 95% confidence intervals (CI) for predicting tau-positivity (defined with tau-PET in WRAP and TRIAD, and with pTau181 in SPIN). The Akaike information criterion (AIC) is also shown.

**Supplementary Table 6.** Binary reference for Aβ-positivity

|  | Binary reference-point for Aβ-positivity,  Plasma pTau217 > 0.42 pg/mL | | |
| --- | --- | --- | --- |
|  | WRAP | TRIAD | SPIN |
| Number of participants | 323 | 268 | 195 |
| Aβ-positive, n (%) | 64 (19.8) | 120 (44.8) | 110 (56.4) |
| Plasma pTau217 status positive, n (%) | 127 (39.3) | 124 (46.3) | 127 (65.1) |
| Sensitivity, % | 95.3 | 85.0 | 98.2 |
| Specificity, % | 74.5 | 85.1 | 77.6 |
| PPA, % | 48.0 | 82.3 | 85.0 |
| NPA, % | 98.5 | 87.5 | 97.1 |
| OPA, % | 78.6 | 85.1 | 89.2 |

The table shows key metrics for the evaluation of a binary ALZpath pTau217 reference-point for Aβ-positivity, derived in WRAP based in the Youden index, and directly cross-validated in TRIAD and SPIN. In WRAP and TRIAD, Aβ-positivity was determined with Aβ-PET, whereas in SPIN with CSF Aβ42/Aβ40. PPA = positive percent agreement. NPA = negative percent agreement. OPA = overall percent agreement.

**Supplementary Table 7.** Three-range reference range for Aβ-positivity

|  | Three-range reference-point for Aβ-positivity,  Plasma pTau217 positive >0.63 pg/mL  Plasma pTau217 negative <0.40 pg/mL | | |
| --- | --- | --- | --- |
|  | WRAP | TRIAD | SPIN |
| Number of participants | 323 | 268 | 195 |
| Aβ-positive, n (%) | 64 (19.8) | 120 (44.8) | 110 (56.4) |
| Plasma pTau217 positive, n (%) | 58 (18.0%) | 86 (32.1%) | 106 (54.4%) |
| Plasma pTau217 intermediate, n (%) | 74 (22.9%) | 43 (16.0%) | 24 (12.3%) |
| Plasma pTau217 negative, n (%) | 191 (59.1%) | 139 (51.9%) | 65 (33.3%) |
| Sensitivity of lower reference-point, % | 95.3 | 86.7 | 98.2 |
| Specificity of upper reference-point, % | 94.9 | 98.6 | 94.1 |
| PPA, upper reference-point, % | 77.6 | 97.7 | 95.3 |
| NPA, lower reference-point, % | 98.4 | 88.5 | 96.9 |
| OPA for pTau217 positive and negative, % | 93.6 | 92.0 | 95.9 |

The table shows key metrics for the evaluation of three-range ALZpath pTau217 reference ranges for Aβ-positivity, derived in WRAP based on 95% sensitivity (lower reference-point) and 95% specificity (upper reference-point), and directly cross-validated in TRIAD and SPIN. In WRAP and TRIAD, Aβ-positivity was determined with Aβ-PET, whereas in SPIN with CSF Aβ42/Aβ40. The OPA for pTau217 negative and positive indicates the combined NPA of those below the lower reference-point and the PPA for those above the upper reference-point, not accounting for the intermediate zone. PPA = positive percent agreement. NPA = negative percent agreement. OPA = overall percent agreement.

**Supplementary Table 8.** Three-range reference range for tau-positivity

|  | Binary reference-point for tau-positivity,  Plasma pTau217 > 0.64 pg/mL | | |
| --- | --- | --- | --- |
|  | WRAP | TRIAD | SPIN |
| Number of participants | 266 | 266 | 184 |
| Tau positive, n (%) | 19 (7.1) | 55 (20.7) | 103 (56.0) |
| Plasma pTau217 positive, n (%) | 49 (18.4) | 82 (30.8) | 99 (53.8) |
| Sensitivity, % | 89.5 | 89.1 | 93.2 |
| Specificity, % | 87.0 | 84.4 | 96.3 |
| PPV, % | 34.7 | 59.8 | 97.0 |
| NPV, % | 99.1 | 96.7 | 91.8 |
| Classification accuracy | 87.0 | 85.3 | 94.6 |

The table shows key metrics for the evaluation of a binary ALZpath pTau217 reference-point for tau-positivity, derived in WRAP based in the Youden index, and directly cross-validated in TRIAD and SPIN. In WRAP and TRIAD, tau-positivity was determined with tau-PET, whereas in SPIN with CSF pTau181. PPA = positive percent agreement. NPA = negative percent agreement. OPA = overall percent agreement.
